# Effect of Paxlovid Treatment During Acute Covid-19 on Long Covid Onset: An EHR-Based Target Trial Emulation from the N3C and RECOVER Consortia

**DOI:** 10.1101/2024.01.20.24301525

**Authors:** Alexander Preiss, Abhishek Bhatia, Leyna V. Aragon, John M. Baratta, Monika Baskaran, Frank Blancero, M. Daniel Brannock, Robert F. Chew, Iván Díaz, Megan Fitzgerald, Elizabeth P. Kelly, Andrea Zhou, Thomas W. Carton, Christopher G. Chute, Melissa Haendel, Richard Moffitt, Emily Pfaff, N3C Consortium and the RECOVER EHR Cohort

**Affiliations:** RTI International, Durham, NC, USA; University of North Carolina at Chapel Hill, Chapel Hill, NC, USA; University of New Mexico, Albuquerque, NM, USA; RECOVER Community Representative; New York University Grossman School of Medicine, New York, NY, USA; Patient Led Research Collaborative; University of Virginia, Charlottesville, VA, USA; Tulane University School of Public Health and Tropical Medicine, New Orleans, LA, USA; Johns Hopkins University School of Medicine, Public Health, and Nursing, Baltimore, MD, USA; Emory University School of Medicine, Atlanta, GA, USA

## Abstract

**Background:** Preventing and treating post-acute sequelae of SARS-CoV-2 infection (PASC), commonly known as Long COVID, has become a public health priority. Researchers have begun to explore whether Paxlovid treatment in the acute phase of COVID-19 could help prevent the onset of PASC.

**Methods and Findings:** We used electronic health records from the National Clinical Cohort Collaborative (N3C) to define a cohort of 410,026 patients who had COVID-19 since April 1, 2022, and were eligible for Paxlovid treatment due to risk for progression to severe COVID-19. We used the target trial emulation framework to estimate the effect of Paxlovid treatment on PASC incidence. The treatment group was defined as outpatients prescribed Paxlovid within five days of COVID-19 index, and the control group was defined as all patients meeting eligibility criteria not in the treatment group. The follow-up period was 180 days. We estimated overall PASC incidence using a computable phenotype. We also measured incident cognitive, fatigue, and respiratory symptoms in the post-acute period. Paxlovid treatment had a small effect on overall PASC incidence (relative risk [RR] 0.94; 95% CI [0.90, 0.99]; p=0.011). It had a slightly stronger protective effect against cognitive (RR 0.86; 95% CI [0.77, 0.95]; p<0.001) and fatigue (RR 0.92; 95% CI [0.86, 0.97]; p=0.002) symptoms.

**Conclusions:** In this study, Paxlovid had a weaker preventative effect on PASC than in prior observational studies, suggesting that Paxlovid is unlikely to become a definitive solution for preventing PASC. Differing effects by symptom cluster suggest that the etiology of cognitive and fatigue symptoms may be more closely related to viral load than that of respiratory symptoms. Future research should explore potential heterogeneous treatment effects across PASC subphenotypes.

**AUTHOR SUMMARY:** *Why was this study done?:*

- Paxlovid is indicated to prevent severe COVID-19.
- Long COVID is more likely after more severe COVID-19, so there is a plausible mechanism for Paxlovid to reduce the risk of developing Long COVID by preventing severe COVID-19.
- Only a few studies have examined the relationship between Paxlovid and Long COVID, with mixed results. To our knowledge, no studies have used causal inference methods to estimate Paxlovid’s effect on PASC.
- If Paxlovid helps prevent Long COVID, it could be a powerful addition to the public health effort to reduce the burden of COVID-19.

*What did the researchers do and find?:*

- We used a cohort of 410,026 patients from the National Covid Cohort Collaborative’s (N3C’s) electronic health record database to estimate the effect of Paxlovid treatment during acute COVID-19 on the likelihood of developing Long COVID.
- We used the target trial emulation technique to estimate the causal effect of Paxlovid treatment using observational data.
- We found that Paxlovid treatment had a small effect on overall Long COVID incidence.
- We found that Paxlovid treatment had a slightly stronger effect on certain symptoms (cognitive and fatigue) of Long COVID.

*What do these findings mean?:*

- Paxlovid is unlikely to become a definitive solution for preventing Long COVID.
- However, Paxlovid may have a stronger effect on certain subtypes of Long COVID.
- This difference in effect by symptom also suggests that viral load may be a more common cause of cognitive and fatigue symptoms than of other Long COVID symptoms.
- This study’s main limitation is that it estimates causal effects, but it is not randomized like a clinical trial. Instead, we control for other variables which could bias the estimate, but if we missed some important variables, the estimates could be incorrect.

## INTRODUCTION

Post-acute sequelae of SARS-CoV-2 infection (PASC), commonly known as Long COVID, affects people from all walks of life. Many people with PASC continue to feel the impacts of the disease years after infection. Mechanisms causing PASC remain largely unknown, and we have yet to identify a treatment that is consistently effective across the array of PASC manifestations. Therefore, developing effective PASC prevention strategies will be crucial to alleviating the long-term public health impact of COVID-19.

Nirmatrelvir with ritonavir (Paxlovid) was given an emergency use authorization (EUA) in the United States in December 2021 for the treatment of patients with mild-to-moderate COVID-19 who are at high risk for progression to severe COVID-19. Paxlovid has proven effective at preventing severe COVID-19, hospitalization, and death, with supporting evidence from clinical trials and real-world evidence.[1–7]

In 2022-23, several teams published case reports where Paxlovid was used to treat extant PASC.[8–11] This evidence motivated several clinical trials, including RECOVER-VITAL, to evaluate Paxlovid as a potential treatment for PASC.[12] Results of smaller trials have begun to emerge. In Stanford University’s STOP-PASC trial, which included 155 participants, Paxlovid did not show benefit in improving extant fatigue, brain fog, body aches, cardiovascular symptoms, shortness of breath, or gastrointestinal symptoms.[13]

In addition to treating PASC, researchers have begun to explore whether Paxlovid treatment in the acute phase of COVID-19 could help prevent the onset of PASC. One plausible pathway could be reducing infection severity. Several studies have found that more severe acute infection or hospitalization is associated with a higher risk of PASC.[14–17] To our knowledge, few studies have explored Paxlovid as a PASC preventative, and results are mixed. The largest study to date used data from the US Department of Veterans Affairs (VA) and found that Paxlovid treatment during the acute phase of COVID-19 was associated with a lower risk of PASC, with a hazard ratio of 0.74.[18] However, two smaller studies found that Paxlovid treatment was not associated with a reduced risk of PASC.[19,20] Because there is still no consensus definition of PASC, these studies used different outcome measures. In sum, the relationship between Paxlovid treatment and PASC onset remains uncertain.

At the time of writing, the PANORAMIC trial in the United Kingdom and the CanTreatCOVID trial in Canada have completed recruitment for arms which will receive Paxlovid during acute COVID-19.[21,22] The PANORAMIC trial will focus on acute outcomes, but the CanTreatCOVID trial will include follow-up at 90 days and 36 weeks. CanTreatCOVID will provide valuable insight to the relationship between Paxlovid treatment and PASC onset.

Through the National Institute of Health’s National COVID Cohort Collaborative (N3C), and as part of the Researching COVID to Enhance Recovery (RECOVER) Initiative’s EHR data team, we have the opportunity to study Paxlovid as a PASC preventative using a large, nationally sampled cohort and an up-to-date study period consisting mostly of Omicron BA and later subvariant infections.[23,24] This study will provide real-world evidence to complement the findings from CanTreatCOVID and, hopefully, additional future trials. All analyses described here were performed within the secure N3C Data Enclave, which integrates EHR data for 21 million patients from over 230 data partners across the United States. N3C’s methods for data acquisition, ingestion, and harmonization have been reported elsewhere.[23,25,26]

## METHODS

We used the target trial emulation (TTE) framework to estimate the effect of Paxlovid treatment in the acute phase of COVID-19 infection on the cumulative incidence of PASC among a cohort of patients eligible for Paxlovid treatment (i.e., with one or more risk factors for developing severe COVID-19).[27] We hypothesized that Paxlovid treatment reduces the likelihood of PASC incidence. We followed the two-step process for emulating target trials with observational data suggested by Hernán and Robins.[28] First, we articulated the causal question of interest in the form of a hypothetical trial protocol. Second, we emulated each component of this protocol using observational EHR data. We did not register a prospective protocol, but the analytical methods did not differ substantially from those planned. See the methods supplement for details.

The study period spanned April 1, 2022, to August 14, 2023, with an index cutoff date of February 28, 2023 (180 days before the end of the study period). Inclusion criteria were: 1) having a documented COVID-19 index date within the study period (with *index date* defined as either a COVID-19 diagnosis [ICD-10 code U07.1] or a positive SARS-CoV-2 test result), 2) being ≥ 18 years of age at the COVID-19 index date (due to potential differences in clinical characteristics and prescription practices between pediatric and adult patients[29,30]), and 3) having ≥ 1 risk factor for severe COVID-19 per CDC guidelines (age ≥ 50 years or diagnosis of a comorbidity associated with higher risk of severe COVID-19[31]). Exclusion criteria were: 1) being hospitalized on the COVID-19 index date, 2) having prior history of PASC, and 3) being prescribed a drug with a severe interaction with Paxlovid in the 30 days prior to the COVID-19 index.[32] To ensure that data were captured from sites with high fidelity and adequate coverage, we excluded sites with fewer than 500 or 5% of eligible patients treated with Paxlovid during the study period. Patients in the cohort were categorized by their treatment exposure. The treatment group was defined as patients prescribed Paxlovid in the outpatient setting within 5 days of their COVID-19 index. The control group was defined as all other eligible patients.

We measured overall PASC incidence using a machine learning-based computable phenotype model, which gathers data for each patient in overlapping 100-day periods that progress through time, and issues a probability of PASC for each 100-day period.[33] The model was trained to classify whether patients have a U09.9 (“Post COVID-19 Condition”) ICD-10 diagnosis code in each period, based on the patients’ diagnoses during each period. The use of a computable phenotype is rare in the PASC literature. More often, researchers measure PASC using specific PASC diagnoses (U09.9) or they define a set of symptoms that constitute PASC and measure their incidence in the post-acute period. However, both of these measurements have problems that the computable phenotype avoids. PASC diagnoses are rare, and diagnosis is likely driven by access to care, which also affects the likelihood of Paxlovid treatment, leading to potential bias. The computable phenotype can help identify undiagnosed patients who have symptoms similar to those with U09.9 diagnoses. Measuring PASC using a specific set of symptoms can lead to false positives (symptoms with etiologies other than COVID-19) and false negatives (related symptoms not included in the definition). The computable phenotype can learn to avoid these errors. However, PASC is also a heterogeneous condition, so the use of symptom-specific outcomes is an important complement to the computable phenotype.

To measure PASC at a more granular level, we also measured incident PASC symptoms in the cognitive, fatigue, and respiratory clusters proposed by the Global Burden of Disease (GBD) Study (“GBD symptom clusters” henceforth).[34] These clusters were the most frequently reported symptoms in a meta-analysis of Long COVID studies.[34] We estimated the effect of treatment on both individual symptom clusters, and a composite measure of any incident symptom across all clusters.

The estimand was the cumulative incidence of PASC from 29 to 180 days after COVID-19 index. We used inverse probability of treatment (IPT) weighting to emulate random assignment through exchangeability between treatment arms and inverse probability of censoring (IPC) weighting to adjust for informative censoring. The treatment and censoring models included the following pre-treatment covariates: sex, age (binned), race and ethnicity, prior history of individual comorbid conditions captured in the Charlson Comorbidity index, value of the composite Charlson Comorbidity Index (CCI; binned), prior history of conditions associated with risk of severe COVID-19 (as defined by the CDC Paxlovid eligibility criteria[31]), Community Well-Being Index (CWBI; binned), number of visits in the year prior to index (binned), number of hospitalizations in the year prior to index (binned), month of COVID-19 onset, and site of care provision. To estimate the cumulative incidence of PASC, we used Aalen-Johansen estimators, where stabilized, trimmed, and combined IP weights were used as time-fixed weights. We used bootstrapping with 200 iterations to estimate the 95% confidence interval at a two-sided alpha of 0.05.

We followed patients for 180 days following their COVID-19 index date. Follow-up started at day 0 and we started observing the outcome at day 29 to avoid attributing acute symptoms to PASC. Computable phenotype PASC predictions and GBD symptoms between days 0 and 28 were ignored. Censorship was possible from days 0 to 180. We censored patients at the following events: 1) death, 2) last documented visit in the EHR, and 3) 180 days after index (end of study period).

We also conducted two subanalyses. The first used a “VA-like cohort” designed to mirror the study period used in Xie et al. (2023) and the demographics of VA patients.[18] It used an earlier study start date (including the first Omicron wave in early 2022) and only included male patients over 65 years old. The intent of this subanalysis was to minimize the differences between our studies and allow for more direct comparison with previously reported findings on this topic. The second subanalysis included COVID-19 vaccination status as an additional covariate, and was conducted in a subset of sites with high-quality vaccination data. We did not include vaccination as a covariate in the primary analysis, because it is subject to substantial measurement error in most EHRs. The intent of this subanalysis was to assess whether vaccination led to unmeasured confounding in the primary analysis. Finally, we conducted several sensitivity analyses to test sensitivity to estimation methods, inclusion and exclusion criteria, computable phenotype prediction threshold, COVID-19 index definition, and time period of outcome observation. See the methods supplement for details.

## RESULTS

### Patient Characteristics

All results are reported in adherence with the Strengthening the Reporting of Observational Studies in Epidemiology (STROBE) guidelines.[35] The study cohort included 410,026 patients, of whom 118,274 (28.85%) were placed in the treatment group, and 16,750 (4.09%) had PASC (U09.9 diagnosis or computable phenotype prediction over 0.9 from 29 to 180 days after index). Among treated patients, 100,706 (85.15%) were prescribed Paxlovid on the same day as COVID-19 index, and 113,798 (96.22%) were prescribed Paxlovid within one day of COVID-19 index. During the study period, 110 (0.09%) patients treated with Paxlovid and 639 (0.22%) untreated patients died. A total of 4,240 (1.03%) patients had a post-acute symptom in the cognitive symptom cluster, 11,899 (2.90%) patients had a post-acute symptom in the fatigue symptom cluster, and 19,116 (4.66%) had a post-acute symptom in the respiratory symptom cluster. Among patients with a PASC diagnosis or computable phenotype prediction, 7.30% had a post-acute symptom in the cognitive symptom cluster, 20.45% had a post-acute symptom in the fatigue symptom cluster, and 36.08% had a post-acute symptom in the respiratory symptom cluster. A co-occurrence matrix, showing the percentage of patients with each outcome who also had other outcomes, is shown in Supplemental Figure 1. After applying the eligibility criteria to the patient population and study sites, a total of 28 of 36 study sites were retained. The CONSORT flow diagram is shown in Figure 1. The characteristics of all patients during the study period are presented in Table 1, stratified by treatment group. IPT weighting achieved balance across all covariates, as shown in Figure 2. The distribution of stabilized, trimmed, and combined IPT and IPC weights had a median of 0.90 and a standard deviation of 0.40. The target trial protocol and emulation approach are presented in Table 2.

**Figure 1:**
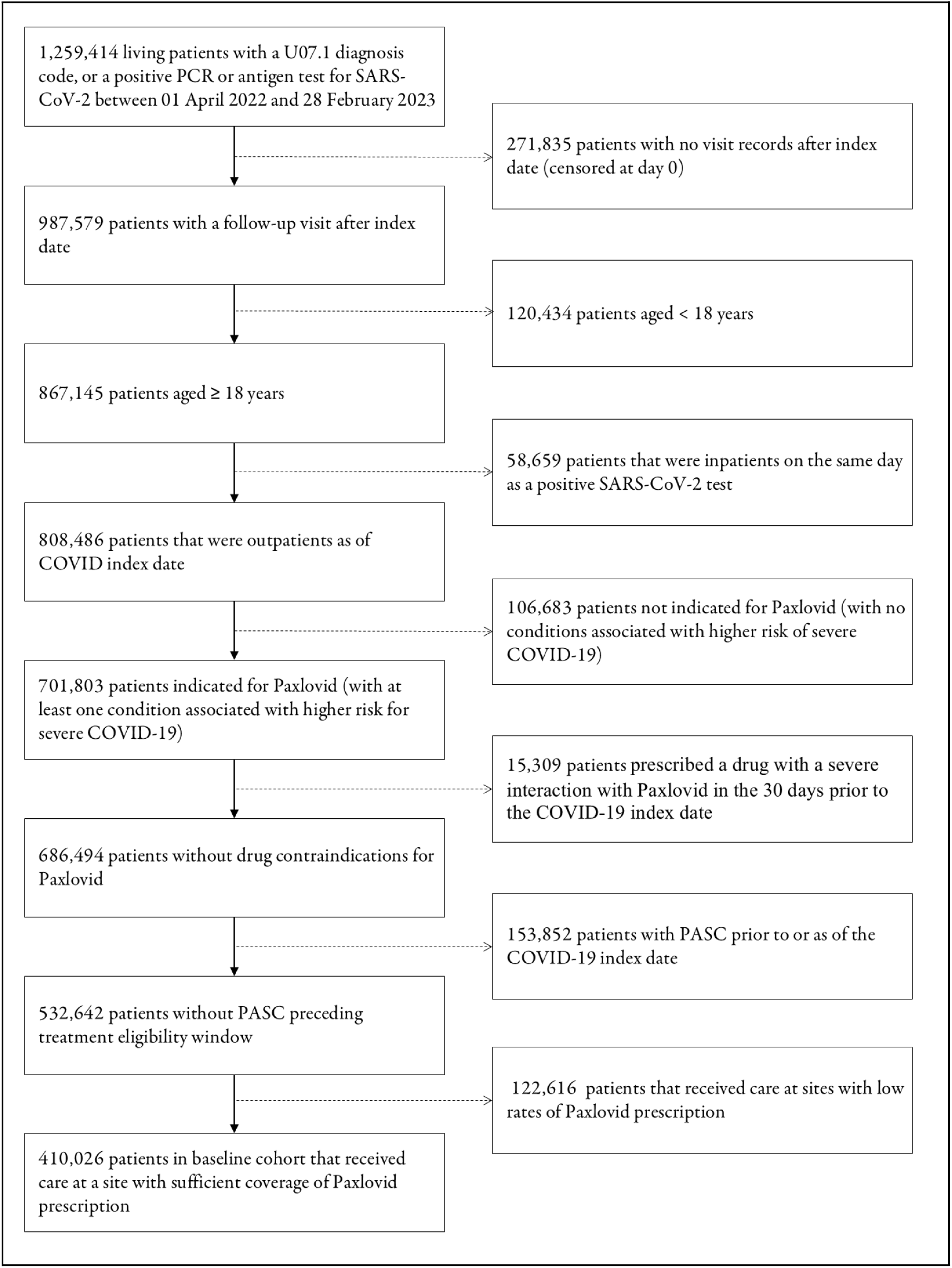
CONSORT Diagram: Study Cohort and Flow of Emulated Trial (PCR: polymerase chain reaction; PASC: post-acute sequelae of COVID-19).

**Figure 2:**
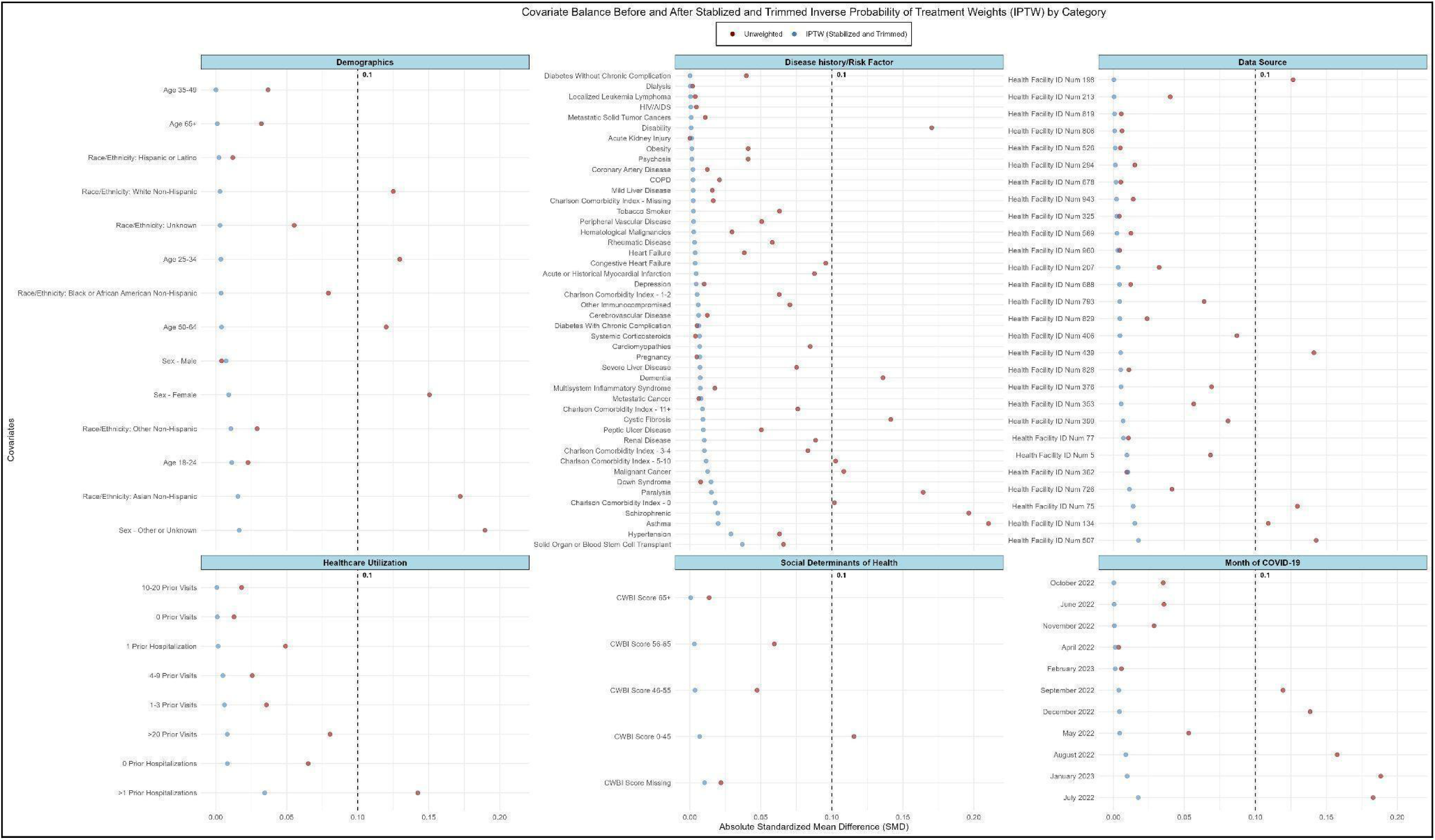
Covariate balance before and after stabilized and trimmed inverse probability of treatment weighting (IPTW).

**Table 1:** Descriptive population characteristics within the N3C Cohort.

| Characteristic | Treatment Group |  |
| --- | --- | --- |
|  | No Paxlovid<br>(N=291752) | Paxlovid<br>(N=118274) |
| PASC |  |  |
| Computable phenotype prediction or diagnosis<sup>1</sup> | 11846 (4.1%) | 4904 (4.1%) |
| Cognitive symptom cluster | 3057 (1.0%) | 1183 (1.0%) |
| Fatigue symptom cluster | 8533 (2.9%) | 3366 (2.8%) |
| Respiratory symptom cluster | 13471 (4.6%) | 5645 (4.8%) |
| Sex |  |  |
| Female | 181831 (62.3%) | 71068 (60.1%) |
| Male | 109880 (37.7%) | 47189 (39.9%) |
| Missing | 41 (0.0%) | <20 (0.0%) |
| Age (in years) |  |  |
| 18-24 | 18959 (6.5%) | 2848 (2.4%) |
| 25-34 | 41133 (14.1%) | 8878 (7.5%) |
| 35-49 | 60133 (20.6%) | 21265 (18.0%) |
| 50-64 | 81474 (27.9%) | 37689 (31.9%) |
| 65+ | 90053 (30.9%) | 47594 (40.2%) |
| Race and Ethnicity |  |  |
| Asian Non-Hispanic | 13338 (4.6%) | 5516 (4.7%) |
| Black or African American Non-Hispanic | 36216 (12.4%) | 11387 (9.6%) |
| Hispanic or Latino Any Race | 31845 (10.9%) | 9494 (8.0%) |
| White Non-Hispanic | 189613 (65.0%) | 84522 (71.5%) |
| Other Non-Hispanic | 5067 (1.7%) | 1336 (1.1%) |
| Unknown | 15673 (5.4%) | 6019 (5.1%) |
| Charlson Comorbidity Index |  |  |
| 0 | 162730 (55.8%) | 65372 (55.3%) |
| 1-2 | 75891 (26.0%) | 36533 (30.9%) |
| 3-4 | 19909 (6.8%) | 8312 (7.0%) |
| 5-10 | 10221 (3.5%) | 3534 (3.0%) |
| 11+ | 732 (0.3%) | 220 (0.2%) |
| Missing | 22269 (7.6%) | 4303 (3.6%) |
| Number of Visits in Prior Year |  |  |
| 0 | 26485 (9.1%) | 6348 (5.4%) |
| 1-3 | 51642 (17.7%) | 14493 (12.3%) |
| 4-9 | 70491 (24.2%) | 28845 (24.4%) |
| 10-20 | 73157 (25.1%) | 36369 (30.7%) |
| > 20 | 69977 (24.0%) | 32219 (27.2%) |
| Number of Hospitalizations in Prior Year |  |  |
| 0 | 278301 (95.4%) | 114533 (96.8%) |
| 1 | 11087 (3.8%) | 3194 (2.7%) |
| > 1 | 2364 (0.8%) | 547 (0.5%) |
| Community Wellbeing Index<sup>2</sup> |  |  |
| 0-45 | 2081 (0.7%) | 618 (0.5%) |
| 46-55 | 105003 (36.0%) | 35399 (29.9%) |
| 56-65 | 132729 (45.5%) | 55923 (47.3%) |
| 65+ | 21627 (7.4%) | 12898 (10.9%) |
| Missing | 30312 (10.4%) | 13436 (11.4%) |
| Censoring Events |  |  |
| Death | 2442 (0.8%) | 420 (0.4%) |
| PASC diagnosis or prediction, day 0 to 28 | 5,145 (1.7%) | 2,151 (1.8%) |
| Lost to follow-up (no further visits in EHR) | 248,316 (23.45%) | 23,518 (11.3%) |
| Month of COVID-19 diagnosis |  |  |
| April 2022 | 17584 (6.0%) | 3643 (3.1%) |
| May 2022 | 40578 (13.9%) | 12480 (10.6%) |
| June 2022 | 41501 (14.2%) | 14268 (12.1%) |
| July 2022 | 44721 (15.3%) | 17846 (15.1%) |
| August 2022 | 37006 (12.7%) | 14454 (12.2%) |
| September 2022 | 23323 (8.0%) | 9606 (8.1%) |
| October 2022 | 18377 (6.3%) | 7094 (6.0%) |
| November 2022 | 17267 (5.9%) | 8217 (6.9%) |
| December 2022 | 23118 (7.9%) | 13978 (11.8%) |
| January 2023 | 17053 (5.8%) | 9471 (8.0%) |
| February 2023 | 11224 (3.8%) | 7217 (6.1%) |

Notes: <sup>1</sup>Any PASC (CP or U09.9) between 28-days following a positive SARS-CoV-2 test result to 180 days post-index; <sup>2</sup>CWBI is a measure of five interrelated community-level domains: Healthcare access (ratios of healthcare providers to population), Resource access (libraries and religious institutions, employment, and grocery stores), Food access (access to grocery stores and produce), Housing & transportation (home values, ratio of home value to income, and public transit use), and Economic security (rates of employment, labor force participation, health insurance coverage rate, and household income above the poverty level).[63]

**Table 2:**
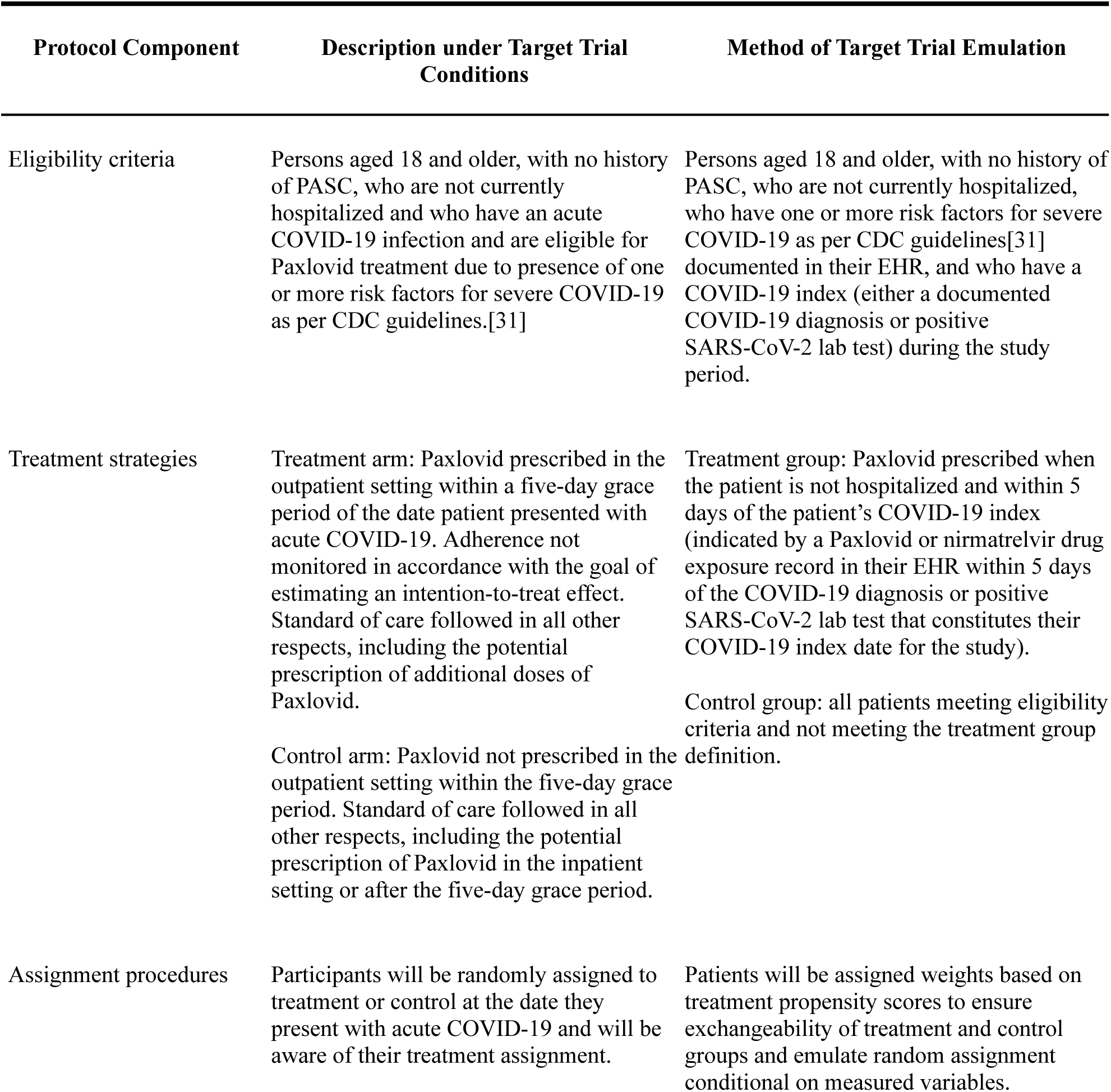
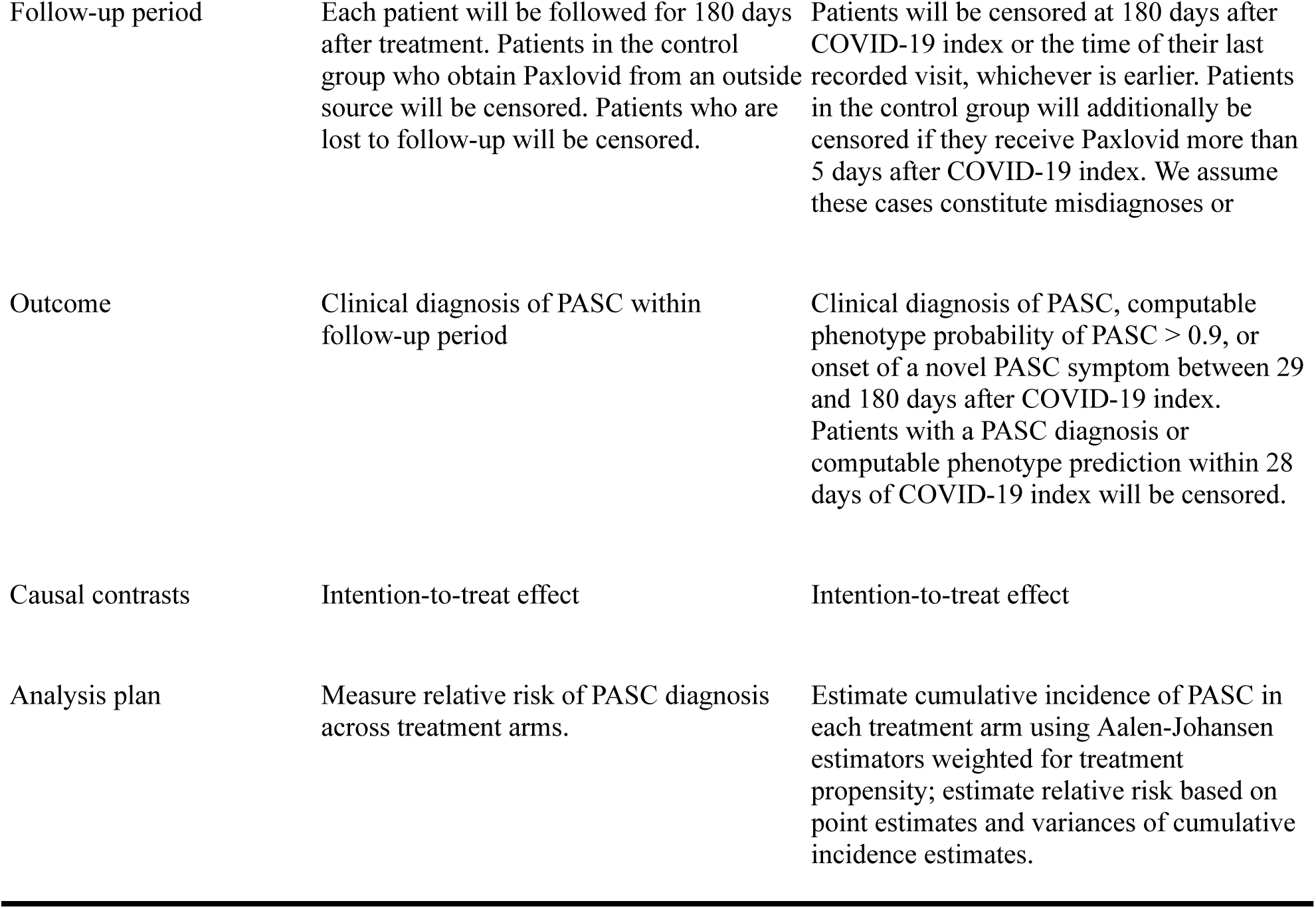
Protocol of a Target Trial Emulation to Estimate the Effect of Paxlovid Treatment during Acute COVID-19 on Cumulative PASC Incidence.

| Protocol Component | Description under Target Trial Conditions | Method of Target Trial Emulation |
| --- | --- | --- |
| Eligibility criteria | Persons aged 18 and older, with no history of PASC, who are not currently hospitalized and who have an acute COVID-19 infection and are eligible for Paxlovid treatment due to presence of one or more risk factors for severe COVID-19 as per CDC guidelines.[31] | Persons aged 18 and older, with no history of PASC, who are not currently hospitalized, who have one or more risk factors for severe COVID-19 as per CDC guidelines[31] documented in their EHR, and who have a COVID-19 index (either a documented COVID-19 diagnosis or positive SARS-CoV-2 lab test) during the study period. |
| Treatment strategies | Treatment arm: Paxlovid prescribed in the outpatient setting within a five-day grace period of the date patient presented with acute COVID-19. Adherence not monitored in accordance with the goal of estimating an intention-to-treat effect. Standard of care followed in all other respects, including the potential prescription of additional doses of Paxlovid.<br>Control arm: Paxlovid not prescribed in the outpatient setting within the five-day grace period. Standard of care followed in all other respects, including the potential prescription of Paxlovid in the inpatient setting or after the five-day grace period. | Treatment group: Paxlovid prescribed when the patient is not hospitalized and within 5 days of the patient's COVID-19 index (indicated by a Paxlovid or nirmatrelvir drug exposure record in their EHR within 5 days of the COVID-19 diagnosis or positive SARS-CoV-2 lab test that constitutes their COVID-19 index date for the study).<br>Control group: all patients meeting eligibility criteria and not meeting the treatment group definition. |
| Assignment procedures | Participants will be randomly assigned to treatment or control at the date they present with acute COVID-19 and will be aware of their treatment assignment. | Patients will be assigned weights based on treatment propensity scores to ensure exchangeability of treatment and control groups and emulate random assignment conditional on measured variables. |
| Follow-up period | Each patient will be followed for 180 days after treatment. Patients in the control group who obtain Paxlovid from an outside source will be censored. Patients who are lost to follow-up will be censored. | Patients will be censored at 180 days after COVID-19 index or the time of their last recorded visit, whichever is earlier. Patients in the control group will additionally be censored if they receive Paxlovid more than 5 days after COVID-19 index. We assume these cases constitute misdiagnoses or |
| Outcome | Clinical diagnosis of PASC within follow-up period | Clinical diagnosis of PASC, computable phenotype probability of PASC $> 0.9$, or onset of a novel PASC symptom between 29 and 180 days after COVID-19 index. Patients with a PASC diagnosis or computable phenotype prediction within 28 days of COVID-19 index will be censored. |
| Causal contrasts | Intention-to-treat effect | Intention-to-treat effect |
| Analysis plan | Measure relative risk of PASC diagnosis across treatment arms. | Estimate cumulative incidence of PASC in each treatment arm using Aalen-Johansen estimators weighted for treatment propensity; estimate relative risk based on point estimates and variances of cumulative incidence estimates. |

### Effect of Paxlovid Treatment on PASC Incidence

Overall, we found that Paxlovid treatment during acute COVID-19 had a small effect on overall PASC onset as defined by our computable phenotype, and a slightly larger effect on cognitive and fatigue symptoms. Table 3 shows estimates of cumulative PASC incidence and treatment effects (relative risk [RR], absolute risk difference [ARD], and number needed to treat [NNT]) across all analyses. Figure 3 shows relative risk for all analyses. Figure 4 shows cumulative incidence functions for the main analyses.

**Figure 3:**
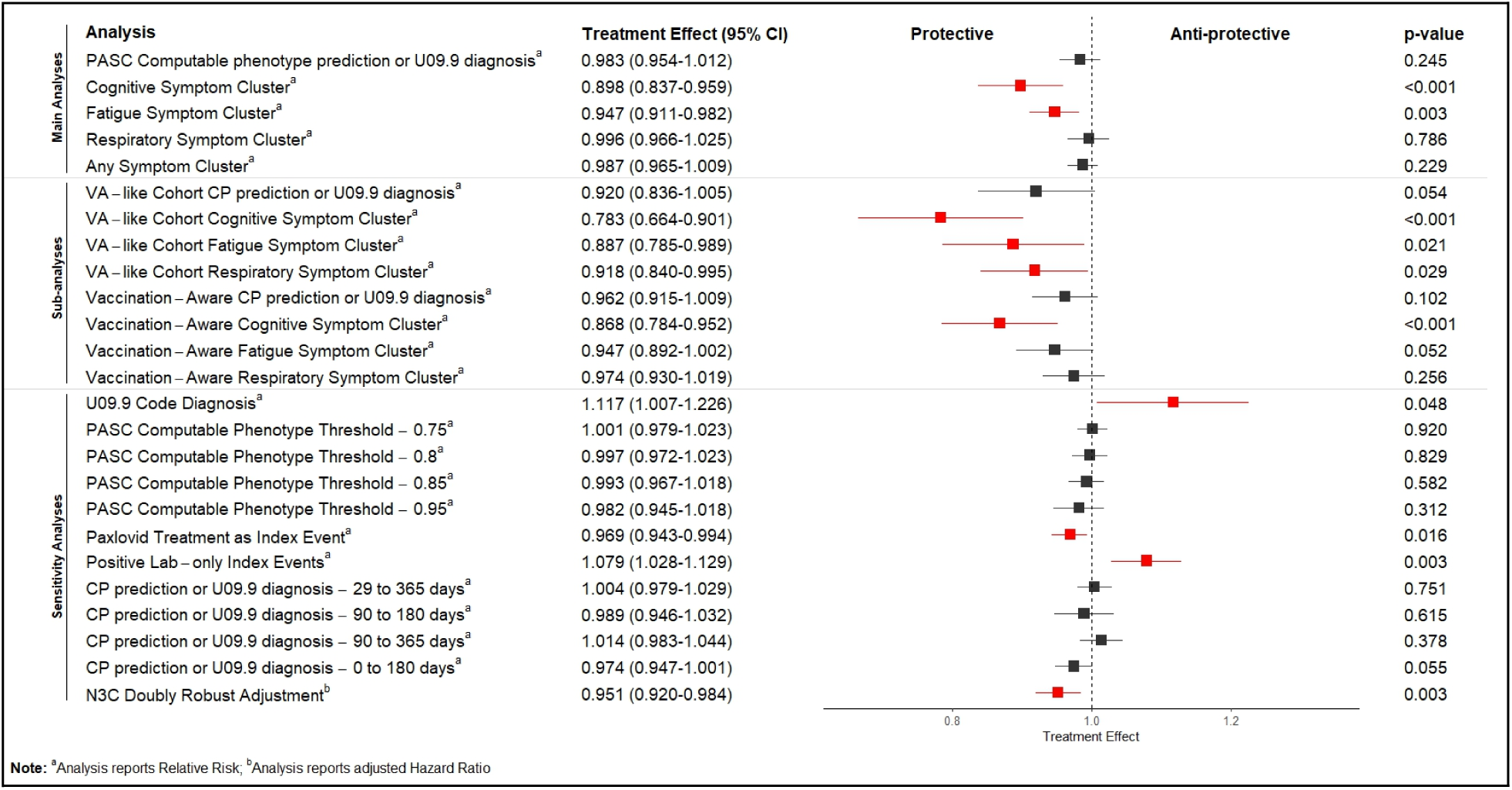
Estimated Treatment Effects (Risk Ratios) of Paxlovid on PASC, across all analyses (PASC: post-acute sequelae of COVID-19; VA: United States Department of Veterans Affairs; CP: computable phenotype).

**Figure 4:**
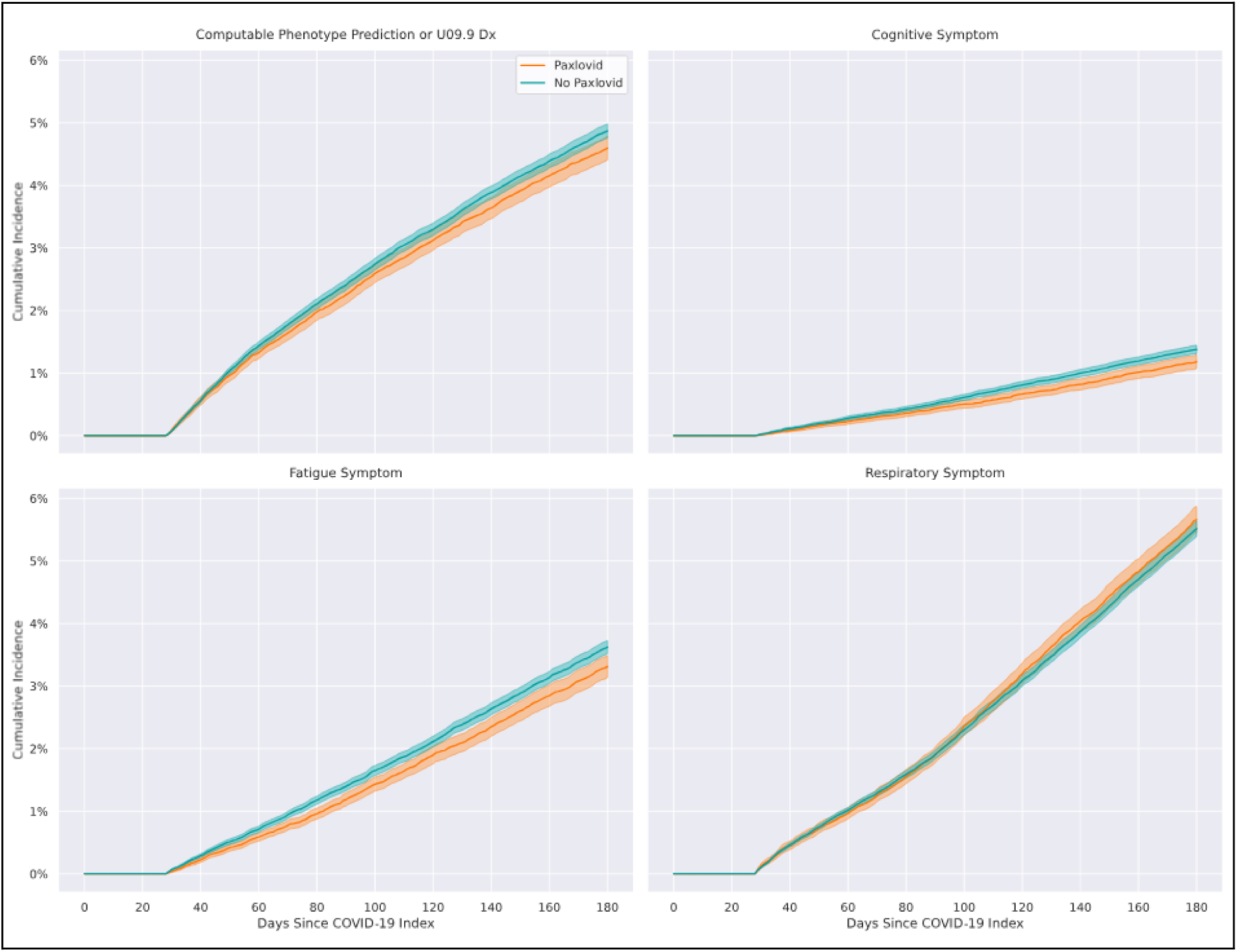
Cumulative incidence of PASC in Paxlovid treated vs. Non-Paxlovid-Treated patients by outcome measure; between 29-180 days. Any GBD Symptom = any symptom from the cognitive, fatigue, and respiratory PASC symptom clusters proposed in the Global Burden of Disease study.

**Table 3:** Cumulative incidence and Absolute Risk Difference estimates across all analyses.

| Analysis | Cumulative Incidence (95% CI) |  | Relative Risk | p-value | Absolute Risk Difference | p-value | NNT |
| --- | --- | --- | --- | --- | --- | --- | --- |
|  | Paxlovid | No Paxlovid |  |  |  |  |  |
| Main Results |  |  |  |  |  |  |  |
| Computable phenotype PASC prediction or U09.9 diagnosis | 0.046 (0.044-0.048) | 0.049 (0.048-0.050) | 0.944 (0.900-0.988) | 0.011 | 0.003 (0.001-0.005) | 0.015 | 366.4 |
| Cognitive Symptom Cluster | 0.012 (0.011-0.013) | 0.014 (0.013-0.014) | 0.858 (0.769-0.948) | <0.001 | 0.002 (0.001-0.003) | 0.003 | 512.4 |
| Fatigue Symptom Cluster | 0.033 (0.031-0.035) | 0.036 (0.035-0.037) | 0.915 (0.859-0.970) | 0.002 | 0.003 (0.001-0.005) | 0.003 | 323.0 |
| Respiratory Symptom Cluster | 0.057 (0.054-0.059) | 0.055 (0.054-0.056) | 1.027 (0.982-1.071) | 0.249 | -0.001 (-0.004-0.001) | 0.240 | - |
| Any Symptom Cluster | 0.093 (0.090-0.095) | 0.094 (0.093-0.096) | 0.983 (0.950-1.015) | 0.288 | 0.002 (-0.001-0.005) | 0.294 | 608.3 |
| Subanalyses |  |  |  |  |  |  |  |
| VA-like Cohort, CP prediction or U09.9 diagnosis | 0.047 (0.042-0.053) | 0.055 (0.052-0.058) | 0.854 (0.745-0.964) | 0.005 | 0.008 (0.002-0.014) | 0.012 | 124.6 |
| VA-like Cohort, Cognitive Symptom Cluster | 0.023 (0.018-0.027) | 0.025 (0.023-0.027) | 0.908 (0.723-1.093) | 0.304 | 0.002 (-0.002-0.007) | 0.336 | 434.3 |
| VA-like Cohort, Fatigue Symptom Cluster | 0.044 (0.037-0.051) | 0.046 (0.043-0.049) | 0.968 (0.804-1.131) | 0.693 | 0.001 (-0.006-0.009) | 0.699 | 672.2 |
| VA-like Cohort, Respiratory Symptom Cluster | 0.065 (0.059-0.071) | 0.067 (0.064-0.070) | 0.971 (0.872-1.071) | 0.565 | 0.002 (-0.005-0.009) | 0.573 | 516.0 |
| Vaccination-Aware, CP prediction or U09.9 diagnosis | 0.042 (0.040-0.044) | 0.046 (0.044-0.048) | 0.913 (0.852-0.973) | 0.003 | 0.004 ( 0.001-0.007) | 0.006 | 249.4 |
| Vaccination-Aware, Cognitive Symptom Cluster | 0.011 (0.009-0.012) | 0.013 (0.012-0.014) | 0.826 (0.707-0.945) | 0.002 | 0.002 ( 0.001-0.004) | 0.007 | 442.1 |
| Vaccination-Aware, Fatigue Symptom Cluster | 0.031 (0.029-0.033) | 0.034 (0.033-0.036) | 0.902 (0.829-0.976) | 0.006 | 0.003 ( 0.001-0.006) | 0.012 | 300.3 |
| Vaccination-Aware, Respiratory Symptom Cluster | 0.047 (0.045-0.050) | 0.047 (0.046-0.049) | 1.003 (0.936-1.071) | 0.923 | 0.000 (-0.003-0.003) | 0.923 | - |
| Sensitivity Analyses |  |  |  |  |  |  |  |
| U09.9 Code Diagnosis | 0.006 (0.005-0.006) | 0.005 (0.005-0.005) | 1.132 (0.961-1.303) | 0.156 | -0.001 (-0.001-0.000) | 0.121 | - |
| PASC Computable Phenotype Threshold - 0.75 | 0.116 (0.114-0.119) | 0.120 (0.118-0.121) | 0.974 (0.947-1.001) | 0.055 | 0.003 (0.000-0.006) | 0.060 | 322.5 |
| PASC Computable Phenotype Threshold - 0.80 | 0.092 (0.090-0.095) | 0.095 (0.094-0.097) | 0.966 (0.937-0.996) | 0.022 | 0.003 (0.000-0.006) | 0.025 | 309.2 |
| PASC Computable Phenotype Threshold - 0.85 | 0.069 (0.067-0.071) | 0.072 (0.071-0.073) | 0.962 (0.929-0.995) | 0.020 | 0.003 (0.000-0.005) | 0.023 | 364.0 |
| PASC Computable Phenotype Threshold - 0.95 | 0.026 (0.024-0.027) | 0.027 (0.026-0.028) | 0.953 (0.890-1.016) | 0.132 | 0.001 (0.000-0.003) | 0.146 | 781.4 |
| Paxlovid Treatment as Index Event | 0.045 (0.044-0.047) | 0.049 (0.048-0.050) | 0.931 (0.890-0.973) | <0.001 | 0.003 (0.001-0.005) | 0.001 | 298.3 |
| Positive Lab-only Index Events | 0.046 (0.045-0.048) | 0.048 (0.045-0.051) | 1.038 (0.967-1.108) | 0.307 | -0.002 (-0.005-0.002) | 0.295 | 574.8 |
| CP prediction or U09.9 diagnosis (29-365 days) | 0.048 (0.046-0.049) | 0.049 (0.049-0.050) | 0.970 (0.935-1.004) | 0.083 | 0.001 (0.000-0.003) | 0.090 | 674.1 |
| CP prediction or U09.9 diagnosis (90-180 days) | 0.035 (0.034-0.037) | 0.037 (0.036-0.037) | 0.969 (0.926-1.013) | 0.157 | 0.001 (0.000-0.003) | 0.167 | 885.3 |
| CP prediction or U09.9 diagnosis (90-365 days) | 0.035 (0.034-0.037) | 0.037 (0.036-0.037) | 0.969 (0.926-1.012) | 0.154 | 0.001 (0.000-0.003) | 0.163 | 885.3 |
| No censoring at last visit date | 0.041 (0.040-0.043) | 0.042 (0.041-0.043) | 0.988 (0.951-1.026) | 0.540 | 0.000 (-0.001-0.002) | 0.544 | 2055.7 |
| Exclude patients with other treatments | 0.047 (0.046-0.049) | 0.049 (0.048-0.050) | 0.973 (0.935-1.010) | 0.151 | 0.001 (-0.001-0.003) | 0.160 | 758.7 |
| Doubly Robust Adjustment | Hazard Ratio: 0.951 (0.920, 0.984) |  |  |  |  |  |  |

For overall PASC onset, measured by our PASC computable phenotype, adjusted cumulative incidence estimates were 4.59% (95% CI [4.41, 4.78]) for treated patients and 4.87% (95% CI [4.76, 4.98]) for untreated patients. The RR of PASC was 0.94 (95% CI [0.90, 0.99]; p=0.011), with an ARD of 0.3% (95% CI [0.1%, 0.5%]; p=0.015) and NNT of 366.4. The RR of any GBD symptom was 0.98 (95% CI [0.95, 1.02]; p=0.288), with an ARD of 0.2% (95% CI [-0.1%, 0.5%]; p=0.294). For the cognitive symptom cluster, RR was 0.86 (95% CI [0.77, 0.95]; p<0.001), with an ARD of 0.2% (95% CI [0.1%, 0.3%]; p=0.003) and NNT of 512.4. For the fatigue symptom cluster, RR was 0.92 (95% CI [0.86, 0.97]; p=0.002), with an ARD of 0.3% (95% CI [0.1%, 0.5%]; p=0.003) and NNT of 323.0. For the respiratory symptom cluster, RR was 1.03 (95% CI [0.98, 1.07]; p=0.249), with an ARD of −0.1% (95% CI [-0.4%, 0.1%]; p=0.240).

### Subanalyses

The VA-like cohort included 65,331 male patients 65 years or older with a COVID-19 index between January 3, 2022, and December 31, 2022.[18] Of this cohort, 17,935 (27.45%) were treated with Paxlovid. RR for PASC overall was 0.85 (95% CI [0.75, 0.96]; p=0.005). For the cognitive, fatigue, and respiratory symptom clusters, RR was 0.91 (95% CI [0.72, 1.09]; p=0.304), 0.97 (95% CI [0.80, 1.13]; p=0.693), and 0.97 (95% CI [0.87, 1.07]; p=0.565), respectively. Cumulative incidence functions are shown in Supplemental Figure 2.

The vaccination-aware cohort included 159,376 patients from 8 sites that met vaccination data quality criteria. Of this cohort, 123,183 (77.3%) were fully vaccinated prior to index, 57,078 (35.81%) were treated with Paxlovid, and 6,265 (3.93%) had PASC. Among fully vaccinated patients, 48,596 (39.5%) were treated, as compared to 8,482 (23.4%) among those not fully vaccinated. RR for PASC overall was 0.91 (95% CI [0.85, 0.97]; p=0.003). For the cognitive, fatigue, and respiratory symptom clusters, RR was 0.83 (95% CI [0.71, 0.95]; p=0.002), 0.90 (95% CI [0.83, 0.98]; p=0.006), and 1.00 (95% CI [0.94, 1.07]; p=0.923), respectively. Cumulative incidence functions are shown in Supplemental Figure 3.

Our findings were not sensitive to adjusting the definition of COVID-19 index events (excluding U07.1 diagnoses without an accompanying lab test or including Paxlovid prescriptions without an accompanying U07.1 diagnosis or lab test), treating death as a competing risk instead of a censoring event, or using a doubly robust estimator and the hazard ratio (HR) of Paxlovid treatment as the estimand. The remaining sensitivity analyses produced slightly smaller effect sizes, with RRs ranging from 0.95 to 0.99. These analyses included varying the computable phenotype prediction threshold, varying the period of PASC observation, assuming that patients did not get PASC during the period after their last documented visit (i.e., not censoring on last visit date), and excluding patients who received Molnupiravir or Ritonavir during the 5-day treatment grace period.

## DISCUSSION

In this target trial emulation using the N3C database and a nationally sampled cohort of patients eligible for Paxlovid treatment (i.e., with one or more risk factors for severe COVID-19), we found that Paxlovid treatment during the acute phase of COVID-19 had a negligible effect on overall PASC incidence (RR 0.94, ARD 0.3%). However, Paxlovid’s effect may be heterogeneous across PASC symptoms. We found that Paxlovid had a slightly stronger protective effect against the onset of novel cognitive (RR 0.86) and fatigue (RR 0.92) symptoms in the post-acute period, but no effect on respiratory symptoms (RR 1.03). In terms of absolute risk reduction, Paxlovid’s effect was no greater for cognitive (ARD 0.2%) or fatigue (ARD 0.3%) symptoms, but we interpret these findings with caution, because the rate at which PASC is documented in the EHR may vary by symptom type. In particular, cognitive symptoms were rare in this study (1.3% overall incidence) but are commonly reported in surveys of patients with PASC[36].

Differing effects by symptom cluster also suggest that Paxlovid may have more impact on the underlying causes of certain symptoms. In the literature, multiple PASC etiologies have been proposed. The chief hypotheses are that, relative to healthy convalescents, those with PASC may be experiencing (1) an aberrant autoimmune response triggered by the virus, (2) organ, tissue, or vascular dysfunction related to inflammatory cascades following infection, and/or (3) persistent viremia due to increased viral load or viral reservoirs. We do not yet know which symptoms are caused by which mechanisms. Paxlovid treatment decreases viral load, and thus could plausibly have more impact on symptoms arising from the third factor.[37] Our findings allow us to generate the hypothesis that viral load may be a more common cause of cognitive and fatigue symptoms than of other PASC symptoms.

In the VA-like subanalysis, Paxlovid’s effect on overall PASC incidence (RR 0.85) was stronger than in the primary analysis (RR 0.94), but it was weaker than the association found in Xie et al. (HR 0.74).[18] Two of our symptom cluster outcomes share many ICD-10 codes with PASC components measured in Xie et al. First, our respiratory symptom cluster (RR 0.97, unadjusted 0.94) aligns with their “shortness of breath” component (HR 0.89, unadjusted RR 0.82). Second, our fatigue symptom cluster (RR 0.97, unadjusted 0.87) aligns with their “fatigue and malaise” component (HR 0.79, unadjusted RR 0.70). Together, these findings suggest that cohort differences explain much of the difference between our primary findings and those of Xie et al. For PASC overall, aligning our cohort on sex, age, and study period accounts for about half the difference in findings. For the symptom clusters, unadjusted RRs are very different from nearly identical PASC components used by Xie et al., which suggests that different statistical methods do not explain the difference in findings. Other aspects of a true VA cohort may explain the remaining difference. Veterans are more likely than demographically similar non-veterans to have been exposed to traumatic brain injury, post-traumatic stress disorder, biohazards, and other risk factors.[38–42] Through consistent access to the VA, the EHR for veterans may also be more complete.[43] Veterans may also differ from demographically similar non-veterans in their access to care.

In the vaccination-aware subanalysis, Paxlovid’s effect on all outcomes was slightly stronger than in the primary analysis. However, we do not interpret this to imply that vaccination caused unmeasured confounding in the primary analysis. Because vaccination is associated with a higher likelihood of getting Paxlovid and a lower likelihood of getting PASC[44], we would expect any confounding to be in the opposite direction (i.e., it would cause an overestimate of Paxlovid’s effect on PASC). Sites in the vaccination-aware subanalysis differed from others in terms of demographics, comorbidity rates, and overall PASC rates, so this finding is likely due to different patient populations rather than unmeasured confounding in the primary analysis.

This study has several strengths that underscore the value of large-scale EHR repositories. We used a large, nationally sampled cohort from 28 sites across the United States, increasing generalizability and decreasing the potential for misclassification present in administrative or claims data.[45] The volume of data in the N3C database allowed for the aggressive inclusion/exclusion criteria necessary for TTE while preserving statistical power.[46,47] Our use of the TTE framework with IPTW and IPCW allowed us to account for confounding and informative censoring and to estimate the causal effect of Paxlovid treatment using observational data.[48–51] Our use of a PASC computable phenotype is also a strength, as described in the Methods section. Finally, the study period makes our findings more relevant than prior studies of this topic, which have included cases from the initial Omicron wave, when Paxlovid was less available and disease dynamics were markedly different.

Three groups of limitations affected this study: 1) limitations related to the cohort definition; 2) limitations related to measurement; and 3) limitations related to observational studies.

This study’s eligibility criteria included eligibility for on-label Paxlovid treatment (i.e., at risk for developing severe COVID-19 due to the presence of one or more risk factors). Therefore, results can only be generalized to a high-risk population. Ideally, a clinical trial of Paxlovid as a PASC preventative would also assess treatment among lower-risk populations. We chose not to emulate such a trial because it would complicate the study design and make exchangeability harder to establish due to confounding by indication. We note that the CanTreatCOVID trial also includes high-risk patients only. The effect of Paxlovid treatment on PASC onset among lower-risk patients is an important area for future research. Additionally, this study’s inclusion criterion of Paxlovid treatment within five days of COVID-19 index differed from the indication of treatment within five days of symptom onset. However, we note that within our cohort, 96% of patients in the treatment group were prescribed Paxlovid within one day of COVID-19 index.

Several variables in this study were subject to measurement error. Many COVID-19 infections during this period were not documented due to the prevalence of home testing, and patients with a documented COVID-19 infection may not be representative of all infected patients. Paxlovid prescriptions from providers outside N3C data partner systems may not be documented. The PASC computable phenotype may also misclassify patients.[33] For this reason, the confidence intervals around computable phenotype-based incidence estimates are likely too narrow. In addition to measurement error, different measures of PASC may account for some of the differences between studies on this topic. Although several institutions have proposed definitions of PASC, they disagree on the symptoms and timing that constitute the condition.[52–55] Finally, this study is subject to limitations common to EHR-based studies. EHRs are susceptible to missing data, and our estimates may be biased if missingness was informative.[56–58]

This study is also subject to the assumptions of all causal inference studies, in particular, that there is no unmeasured confounding. One potential unmeasured confounder is acute COVID-19 severity prior to index. Sicker patients may be more likely to seek Paxlovid and develop PASC. The EHR contains no reliable measure of this construct, but we control for pre-diagnosis comorbidities, which have been shown to correlate so strongly with COVID-19 severity that they can be considered proxies, thus mitigating the potential unmeasured confounding from this source.[59–61] Propensity to seek healthcare and access to care may be additional unmeasured confounders, but we control for utilization in the prior year as a proxy for these constructs.

In conclusion, we see Paxlovid as unlikely to become a definitive solution for PASC prevention. Although it had a small protective effect, an absolute risk reduction of 0.3% means that over 300 people would need to be treated with Paxlovid to prevent one case of PASC. Paxlovid remains an important tool to reduce the pandemic’s public health burden by preventing hospitalization and death due to acute COVID-19. Its stronger protective effect against cognitive and fatigue symptoms also merits further research on potential heterogeneous treatment effects across PASC subphenotypes. However, broadly effective interventions to prevent PASC remain elusive.

## ACKNOWLEDGEMENTS

This study is part of the NIH Researching COVID to Enhance Recovery (RECOVER) Initiative, which seeks to understand, treat, and prevent the post-acute sequelae of SARS-CoV-2 infection (PASC). For more information on RECOVER, visit https://recovercovid.org/. This research was funded by the National Institutes of Health (NIH) Agreement OTA OT2HL161847 as part of the Researching COVID to Enhance Recovery (RECOVER) research program.

The analyses described in this manuscript were conducted with data or tools accessed through the NCATS N3C Data Enclave https://covid.cd2h.org and N3C Attribution & Publication Policy v 1.2-2020-08-25b supported by NCATS U24 TR002306, Axle Informatics Subcontract: NCATS-P00438-B, and by the RECOVER Initiative (OT2HL161847–01). The N3C Publication committee confirmed that this manuscript msid:1733.497 is in accordance with N3C data use and attribution policies; however, this content is solely the responsibility of the authors and does not necessarily represent the official views of the National Institutes of Health or the RECOVER or N3C programs. This research was possible because of the patients whose information is included within the data and the organizations (https://ncats.nih.gov/n3c/resources/data-contribution/ data-transfer-agreement-signatories) and scientists who have contributed to the on-going development of this community resource [https://doi.org/10.1093/jamia/ocaa196]. We would also like to thank the National Community Engagement Group (NCEG), all patient, caregiver and community Representatives, and all the participants enrolled in the RECOVER Initiative.

We also acknowledge the following institutions whose data is released or pending:

Available: Advocate Health Care Network — UL1TR002389: The Institute for Translational Medicine (ITM) • Aurora Health Care Inc — UL1TR002373: Wisconsin Network For Health Research • Boston University Medical Campus — UL1TR001430: Boston University Clinical and Translational Science Institute • Brown University — U54GM115677: Advance Clinical Translational Research (Advance-CTR) • Carilion Clinic — UL1TR003015: iTHRIV Integrated Translational health Research Institute of Virginia • Case Western Reserve University — UL1TR002548: The Clinical & Translational Science Collaborative of Cleveland (CTSC) • Charleston Area Medical Center — U54GM104942: West Virginia Clinical and Translational Science Institute (WVCTSI) • Children’s Hospital Colorado — UL1TR002535: Colorado Clinical and Translational Sciences Institute • Columbia University Irving Medical Center — UL1TR001873: Irving Institute for Clinical and Translational Research • Dartmouth College — None (Voluntary) Duke University — UL1TR002553: Duke Clinical and Translational Science Institute • George Washington Children’s Research Institute — UL1TR001876: Clinical and Translational Science Institute at Children’s National (CTSA-CN) • George Washington University — UL1TR001876: Clinical and Translational Science Institute at Children’s National (CTSA-CN) • Harvard Medical School — UL1TR002541: Harvard Catalyst • Indiana University School of Medicine — UL1TR002529: Indiana Clinical and Translational Science Institute • Johns Hopkins University — UL1TR003098: Johns Hopkins Institute for Clinical and Translational Research • Louisiana Public Health Institute — None (Voluntary) • Loyola Medicine — Loyola University Medical Center • Loyola University Medical Center — UL1TR002389: The Institute for Translational Medicine (ITM) • Maine Medical Center — U54GM115516: Northern New England Clinical & Translational Research (NNE-CTR) Network • Mary Hitchcock Memorial Hospital & Dartmouth Hitchcock Clinic — None (Voluntary) • Massachusetts General Brigham — UL1TR002541: Harvard Catalyst • Mayo Clinic Rochester — UL1TR002377: Mayo Clinic Center for Clinical and Translational Science (CCaTS) • Medical University of South Carolina — UL1TR001450: South Carolina Clinical & Translational Research Institute (SCTR) • MITRE Corporation — None (Voluntary) • Montefiore Medical Center — UL1TR002556: Institute for Clinical and Translational Research at Einstein and Montefiore • Nemours — U54GM104941: Delaware CTR ACCEL Program • NorthShore University HealthSystem — UL1TR002389: The Institute for Translational Medicine (ITM) • Northwestern University at Chicago — UL1TR001422: Northwestern University Clinical and Translational Science Institute (NUCATS) • OCHIN — INV-018455: Bill and Melinda Gates Foundation grant to Sage Bionetworks • Oregon Health & Science University — UL1TR002369: Oregon Clinical and Translational Research Institute • Penn State Health Milton S. Hershey Medical Center — UL1TR002014: Penn State Clinical and Translational Science Institute • Rush University Medical Center — UL1TR002389: The Institute for Translational Medicine (ITM) • Rutgers, The State University of New Jersey — UL1TR003017: New Jersey Alliance for Clinical and Translational Science • Stony Brook University — U24TR002306 • The Alliance at the University of Puerto Rico, Medical Sciences Campus — U54GM133807: Hispanic Alliance for Clinical and Translational Research (The Alliance) • The Ohio State University — UL1TR002733: Center for Clinical and Translational Science • The State University of New York at Buffalo — UL1TR001412: Clinical and Translational Science Institute • The University of Chicago — UL1TR002389: The Institute for Translational Medicine (ITM) • The University of Iowa — UL1TR002537: Institute for Clinical and Translational Science • The University of Miami Leonard M. Miller School of Medicine — UL1TR002736: University of Miami Clinical and Translational Science Institute • The University of Michigan at Ann Arbor — UL1TR002240: Michigan Institute for Clinical and Health Research • The University of Texas Health Science Center at Houston — UL1TR003167: Center for Clinical and Translational Sciences (CCTS) • The University of Texas Medical Branch at Galveston — UL1TR001439: The Institute for Translational Sciences • The University of Utah — UL1TR002538: Uhealth Center for Clinical and Translational Science • Tufts Medical Center — UL1TR002544: Tufts Clinical and Translational Science Institute • Tulane University — UL1TR003096: Center for Clinical and Translational Science • The Queens Medical Center — None (Voluntary) • University Medical Center New Orleans — U54GM104940: Louisiana Clinical and Translational Science (LA CaTS) Center • University of Alabama at Birmingham — UL1TR003096: Center for Clinical and Translational Science • University of Arkansas for Medical Sciences — UL1TR003107: UAMS Translational Research Institute • University of Cincinnati — UL1TR001425: Center for Clinical and Translational Science and Training • University of Colorado Denver, Anschutz Medical Campus — UL1TR002535: Colorado Clinical and Translational Sciences Institute • University of Illinois at Chicago — UL1TR002003: UIC Center for Clinical and Translational Science • University of Kansas Medical Center — UL1TR002366: Frontiers: University of Kansas Clinical and Translational Science Institute • University of Kentucky — UL1TR001998: UK Center for Clinical and Translational Science • University of Massachusetts Medical School Worcester — UL1TR001453: The UMass Center for Clinical and Translational Science (UMCCTS) • University Medical Center of Southern Nevada — None (voluntary) • University of Minnesota — UL1TR002494: Clinical and Translational Science Institute • University of Mississippi Medical Center — U54GM115428: Mississippi Center for Clinical and Translational Research (CCTR) • University of Nebraska Medical Center — U54GM115458: Great Plains IDeA-Clinical & Translational Research • University of North Carolina at Chapel Hill — UL1TR002489: North Carolina Translational and Clinical Science Institute • University of Oklahoma Health Sciences Center — U54GM104938: Oklahoma Clinical and Translational Science Institute (OCTSI) • University of Pittsburgh — UL1TR001857: The Clinical and Translational Science Institute (CTSI) • University of Pennsylvania — UL1TR001878: Institute for Translational Medicine and Therapeutics • University of Rochester — UL1TR002001: UR Clinical & Translational Science Institute • University of Southern California — UL1TR001855: The Southern California Clinical and Translational Science Institute (SC CTSI) • University of Vermont — U54GM115516: Northern New England Clinical & Translational Research (NNE-CTR) Network • University of Virginia — UL1TR003015: iTHRIV Integrated Translational health Research Institute of Virginia • University of Washington — UL1TR002319: Institute of Translational Health Sciences • University of Wisconsin-Madison — UL1TR002373: UW Institute for Clinical and Translational Research • Vanderbilt University Medical Center — UL1TR002243: Vanderbilt Institute for Clinical and Translational Research • Virginia Commonwealth University — UL1TR002649: C. Kenneth and Dianne Wright Center for Clinical and Translational Research • Wake Forest University Health Sciences — UL1TR001420: Wake Forest Clinical and Translational Science Institute • Washington University in St. Louis — UL1TR002345: Institute of Clinical and Translational Sciences • Weill Medical College of Cornell University — UL1TR002384: Weill Cornell Medicine Clinical and Translational Science Center • West Virginia University — U54GM104942: West Virginia Clinical and Translational Science Institute (WVCTSI) Submitted: Icahn School of Medicine at Mount Sinai — UL1TR001433: ConduITS Institute for Translational Sciences • The University of Texas Health Science Center at Tyler — UL1TR003167: Center for Clinical and Translational Sciences (CCTS) • University of California, Davis — UL1TR001860: UCDavis Health Clinical and Translational Science Center • University of California, Irvine — UL1TR001414: The UC Irvine Institute for Clinical and Translational Science (ICTS) • University of California, Los Angeles — UL1TR001881: UCLA Clinical Translational Science Institute • University of California, San Diego — UL1TR001442: Altman Clinical and Translational Research Institute • University of California, San Francisco — UL1TR001872: UCSF Clinical and Translational Science Institute Pending: Arkansas Children’s Hospital — UL1TR003107: UAMS Translational Research Institute • Baylor College of Medicine — None (Voluntary) • Children’s Hospital of Philadelphia — UL1TR001878: Institute for Translational Medicine and Therapeutics • Cincinnati Children’s Hospital Medical Center — UL1TR001425: Center for Clinical and Translational Science and Training • Emory University — UL1TR002378: Georgia Clinical and Translational Science Alliance • HonorHealth — None (Voluntary) • Loyola University Chicago — UL1TR002389: The Institute for Translational Medicine (ITM) • Medical College of Wisconsin — UL1TR001436: Clinical and Translational Science Institute of Southeast Wisconsin • MedStar Health Research Institute — None (Voluntary) • Georgetown University — UL1TR001409: The Georgetown-Howard Universities Center for Clinical and Translational Science (GHUCCTS) • MetroHealth — None (Voluntary) • Montana State University — U54GM115371: American Indian/Alaska Native CTR • NYU Langone Medical Center — UL1TR001445: Langone Health’s Clinical and Translational Science Institute • Ochsner Medical Center — U54GM104940: Louisiana Clinical and Translational Science (LA CaTS) Center • Regenstrief Institute — UL1TR002529: Indiana Clinical and Translational Science Institute • Sanford Research — None (Voluntary) • Stanford University — UL1TR003142: Spectrum: The Stanford Center for Clinical and Translational Research and Education • The Rockefeller University — UL1TR001866: Center for Clinical and Translational Science • The Scripps Research Institute — UL1TR002550: Scripps Research Translational Institute • University of Florida — UL1TR001427: UF Clinical and Translational Science Institute • University of New Mexico Health Sciences Center — UL1TR001449: University of New Mexico Clinical and Translational Science Center • University of Texas Health Science Center at San Antonio — UL1TR002645: Institute for Integration of Medicine and Science • Yale New Haven Hospital — UL1TR001863: Yale Center for Clinical Investigation

## AUTHOR CONTRIBUTIONS

Authorship was determined according to ICMJE recommendations.

Alexander Preiss and Abhishek Bhatia contributed to study design, methodology, analysis, writing, and manuscript review.

Leyna Aragon contributed to study design and writing.

John M Baratta contributed to study design and manuscript review.

Monika Baskaran contributed to analysis.

Frank Blancero contributed to study design and writing.

M Daniel Brannock contributed to study design, methodology, writing, and manuscript review. Robert F Chew and Ivan Diaz contributed to study design, methodology, and manuscript review. Elizabeth P Kelly contributed to project management.

Andrea G Zhou contributed to analysis.

Thomas W Carton, Chistopher G Chute, and Melissa Haendel contributed to study design and manuscript review.

Richard Moffitt contributed to study design, methodology, and manuscript review. Emily Pfaff contributed to study design, methodology, writing, and manuscript review.

## Ethics approval and Consent to Participate

The N3C data transfer to NCATS is performed under a Johns Hopkins University Reliance Protocol # IRB00249128 or individual site agreements with NIH. The N3C Data Enclave is managed under the authority of the NIH; information can be found at https://ncats.nih.gov/n3c/resources. The work was performed under DUR RP-5677B5.

## Data Availability

All data is available in the N3C Data Enclave to those with an approved protocol and data use request from an institutional review board. Data access is governed under the authority of the National Institutes of Health; more information on accessing the data can be found at https://covid.cd2h.org/for-researchers. See Haendel et. al. for additional detail on how data is ingested, managed, and protected within the N3C Data Enclave[23].

## Code Availability

The N3C Enclave is available for public use. To access data used within this manuscript, institutions must have a signed Data Use Agreement executed with the U.S. National Center for Advancing Translational Sciences (NCATS) and their investigators must complete mandatory training and must submit a Data Use Request (DUR) to N3C. To request N3C data access, researchers must follow instructions at https://covid.cd2h.org/onboarding. Code is available to those with valid login credentials for the N3C Data Enclave. It was written for use in the enclave on the Palantir Foundry platform[62], where the analysis can be reproduced by researchers. It can be exported for review upon request.

---

**S1 Figure A:** Outcome co-occurrence matrix. Each cell represents the percentage of patients with the row outcome who also had the column outcome.

**S1 Figure B:** Cumulative incidence of PASC in Paxlovid treated vs. Non-Paxlovid-Treated patients by outcome measure; between 29-180 days; VA-like subanalysis

**S1 Figure C:** Cumulative incidence of PASC in Paxlovid treated vs. Non-Paxlovid-Treated patients by predicted outcome from CP model with threshold of 0.9 or U09.9, additionally adjusted for vaccination status and among data partners meeting vaccination data quality criteria

**S1 Figure D:** Causal diagram used to inform covariate selection. Treatment is shown in green; outcome is shown in orange; observed covariates are shown in gray; unobserved covariates are shown in pink. Note that this diagram only shows the relationships relevant to this study. Covariates may have other causes that are omitted for clarity.

**S1 Table A:** ICD-10 codes used to define Global Burden of Disease symptom clusters.

