## Supplementary Material S1 for "Effect of Paxlovid Treatment During Acute Covid-19 on Long Covid Onset: An EHR-Based Target Trial Emulation from the N3C and RECOVER Consortia"

**Extended Methods**

*Study Period*

The study period spanned April 1, 2022, to August 14, 2023, with an index cutoff date of February 28, 2023 (180 days before the end of the study period). We chose not to study the period between December 21, 2021 (date of Paxlovid EUA) and March 31, 2022 due to the variability in case counts and prescription patterns during the first wave of the Omicron variant.[1] During the first Omicron wave, COVID-19 incidence was much higher than later in the study period, and Paxlovid treatment was much rarer. This would make it difficult to meet the positivity assumption required for causal inference, because the large majority of untreated patients would come from the first Omicron wave, and the large majority of treated patients would come from later in the study period.

*Treatment and Outcome*

We chose to emulate a pragmatic target trial, in which patients who were not randomized to receive the treatment of interest could still be treated to the standard of care in all other respects, including the possible prescription of Paxlovid in ways that do not meet the treatment definition. Patients who were prescribed Paxlovid in the inpatient setting within 5 days of COVID-19 index, or Paxlovid in any setting more than 5 days after COVID-19 index, were included in the control group. We took this approach because inpatient Paxlovid treatment (presumably after COVID-19 is already severe) is a different treatment modality, and we intended to study on-label outpatient treatment. We selected a treatment window of 5 days from COVID-19 index to adhere as closely as possible to treatment guidelines (within 5 days of symptom onset) with the available data. We identified 10 Observational Medical Outcomes Partnership [OMOP] concepts that correspond to Paxlovid in N3C and used these concepts to measure treatment.[2]

We considered two measures of the PASC outcome. To measure PASC overall, we used a computable phenotype: a machine learning model trained to predict PASC diagnoses (ICD-10 code U09.9). An earlier version of this computable phenotype was used in prior work.[3] For this study, we used an updated version better suited for the later phase of the pandemic.[3,4] The model gathers data for each patient in overlapping 100-day periods that progress through time, and issues a probability of PASC for each 100-day period. The model was trained to classify whether patients have a U09.9 ("Post COVID-19 Condition") ICD-10 diagnosis code in each period, based on the patients' diagnoses during each period. PASC date was defined as the start date of the 100-day period which had the maximum computable phenotype prediction above a threshold of 0.9, or, if present, the date of U09.9 diagnosis, whichever was earlier. Patients over 100 years old at COVID-19 index did not receive model scores and were excluded from analysis of this outcome.

To measure PASC at a more granular level, we examined the PASC symptom clusters--cognitive, fatigue, and respiratory--proposed by the Global Burden of Disease (GBD) Study.[5] These clusters were the most frequently reported symptoms in a meta-analysis of Long COVID studies. Their full definitions are cognitive problems (forgetfulness or difficulty concentrating, commonly referred to as brain fog); persistent fatigue with bodily pain (myalgia) or mood swings; and ongoing respiratory problems (primarily shortness of breath and persistent cough). For the GBD symptom cluster outcomes, we conducted two distinct types of analyses. First, we examined the effect of treatment on the onset of each GBD symptom cluster independently, with PASC date defined as the first diagnosis date of any incident symptom in the cluster at least 29 days after COVID-19 index (we defined incident symptoms as symptoms that did not occur in the three years prior to COVID-19 index). Second, we examined the effect of treatment on a composite symptom-based outcome, with PASC date defined as the earliest post-acute onset date of any incident symptom in any of the three GBD symptom clusters. The list of ICD-10 codes to define each GBD symptom cluster cluster was based on the GBD study and is presented in Supplemental Table 1. Individuals were assessed for symptom cluster outcomes regardless of their computable phenotype PASC outcomes. The symptom clusters are not mutually exclusive.

A positive prediction from the computable phenotype model does not imply that a patient must have a positive outcome for one or more symptom clusters. The model considers many more diagnosis codes than those included in the symptom clusters (see the "SNOMED Roll Up" section in the supplement of Crosskey et al., 2023), and a positive prediction may be based on other diagnosis codes.[6] Also, the computable phenotype model features are not restricted to incident diagnoses. For example, if a patient had a dyspnea diagnosis in the three years prior to index, a post-acute dyspnea diagnosis would not count for the respiratory symptom cluster, but it would be considered by the computable phenotype model.

*Statistical Analysis*

We used a single logistic regression model to estimate each patient's propensity of treatment based on a set of baseline covariates. We selected covariates based on a theoretical causal model, which the author team - consisting of clinicians, epidemiologists, bioinformaticians, data scientists, and patient representatives with lived experience - developed collaboratively. Our causal model is shown as a directed acyclic graph in Supplemental Figure 4. Our specific rationale for selected covariates is as follows. Many studies have shown disparity in COVID-19 treatment and outcome by race, ethnicity, and social determinants of health.[7–10] Sex, age, and comorbidities are known to affect care seeking and the outcome of COVID-19. Past healthcare utilization could affect the likelihood of treatment seeking and PASC documentation. Finally, the index month was included because Paxlovid treatment rates, viral variants, and infection rates changed during the study period. CCI was coded as missing when no condition records were present in N3C prior to index. CWBI was coded as missing when patient ZIP code was not reported.

We used this treatment model to generate stabilized IPT weights for each individual as $proportion\ treated\ /\ propensity\ score$ for the treatment group and $proportion\ treated\ /\ (1\ -\ propensity\ score)$ for the control group[11]. To reduce the influence of extreme weights, we trimmed IPT weights at the 99.5th percentile. We assessed covariate balance using absolute standardized differences.

We also generated inverse probability of censoring (IPC) weights to adjust for informative censoring. Death and loss to follow-up were more common in the control group, which could lead to bias. IPC weighting produces a pseudo-cohort in which censoring is random with respect to treatment. To generate IPC weights, we used a single logistic regression model to estimate each patient's propensity of censorship during the study period, based on their treatment group and the same set of covariates as the treatment model. This approach treats the likelihood of censoring as time-invariant, an assumption that we verified by examining the cumulative incidence of censoring by treatment group over time. We used this model to generate stabilized

IPC weights as $proportion\ censored\ /\ propensity\ to\ be\ censored$ for censored patients and $proportion\ censored\ /\ (1\ -\ propensity\ to\ be\ censored)$ for uncensored patients[12]. We also trimmed IPC weights at the 99.5th percentile. Finally, we generated combined inverse probability weights as the product of IPT and IPC weights.

By treating death as a censoring event rather than a competing risk, we estimated the direct effect of Paxlovid treatment on PASC incidence, not including any effect that is mediated by death.[13] Paxlovid is known to reduce mortality, and patients who would have died without Paxlovid treatment are at high risk for PASC. Therefore, Paxlovid could essentially convert some deaths into PASC, which would produce an anti-protective component of its effect. However, any such effect is unlikely to affect clinical treatment decisions. Censoring at death removes this effect from the overall effect estimate. Also, to assess the magnitude of this effect, we conducted a sensitivity test in which death was treated as a competing risk.

*Subanalyses*

In the first subanalysis, we attempted to mirror the cohort used in Xie et al (2023) to make our study more comparable to prior knowledge. In this analysis, we used the same study start and end dates as Xie et al. (January 3, 2022, and December 31, 2022). To mirror VA demographics, we filtered the cohort to males >= 65 years old at COVID-19 index.

In the second subanalysis, we included COVID-19 vaccination status as a covariate, and replicated our primary analysis. We considered vaccination to be a plausible confounder of Paxlovid treatment and documented PASC, either through acute infection severity or propensity to seek care. However, vaccination status in N3C (like most EHRs) is subject to missingness. In this subanalysis, we used a subcohort of patients from sites with reliable vaccination data, which we identified, as in prior work, by comparing each site's data to public vaccination rates for its catchment area.[3,14] We categorized patients by their vaccination status prior to their COVID-19 index date, defined as having completed a full course of vaccination at least 14 days prior to index. Partially vaccinated patients and patients who became fully vaccinated fewer than 14 days prior to index were excluded from the analysis.

*Sensitivity Analyses*

We conducted seven sensitivity analyses.

First, we used a doubly-robust estimation method in case the treatment model was misspecified. Computationally expensive doubly-robust methods like targeted maximum likelihood estimation were not feasible with our cohort and computing environment, so we were unable to estimate cumulative incidence using a doubly-robust method. Instead, we estimated the hazard ratio (HR) of Paxlovid treatment as a secondary estimand. We used IPT- and IPC-weighted Cox proportional hazards models adjusted for the same baseline covariates as the treatment model. The same bootstrap procedure was used to estimate confidence intervals.

Second, we tested various computable phenotype prediction thresholds. In addition to the 0.9 threshold used in the primary analysis, we tested prediction thresholds at 0.75, 0.8, 0.85, and 0.95.

Third, we included Paxlovid treatment as a COVID-19 index event. This added 33,008 additional patients who were treated with Paxlovid during the study period, but did not have a U07.1 diagnosis or a positive lab test in the five days prior to treatment.

Fourth, we also tested sensitivity to COVID-19 index definition by including only positive lab tests as index events. This removed 224,148 patients who had U07.1 diagnoses without accompanying lab results.

Fifth, we tested sensitivity to outcome definition in three ways: by requiring outcomes to occur 90 days after COVID-19 index (rather than 29 days), by observing patients for up to 365 days (rather than 180 days), and by the combination of both (observing PASC from days 90 to 365).

Sixth, we tested sensitivity to our censoring approach. In one analysis, we treated death as a competing risk rather than a censoring event. We used IP-weighted Kaplan-Meier models to estimate the overall survival function (for death or PASC) and the cause-specific hazard function (for PASC). We computed the cause-specific incidence function as $cause\ specific\ hazard\ *\ overall\ survival$. This includes any effect mediated by death in the overall estimate of Paxlovid's effect on PASC. In another analysis, we did not censor patients at the date of their last documented visit in the EHR. This assumes that patients did not get care in the time after their last visit, rather than assuming that they were lost to follow-up and could have gotten care elsewhere.

Seventh, we tested sensitivity to the treatment and control arm definitions in our target trial. In the primary analysis, the target trial is pragmatic, and does not exclude patients who received other treatments. In this sensitivity analysis, we excluded patients who received two other COVID-19 treatments - Molnupiravir and Ritonavir - from both study arms.

**S1 Figure A: Outcome co-occurrence matrix. Each cell represents the percentage of patients with the row outcome who also had the column outcome.**

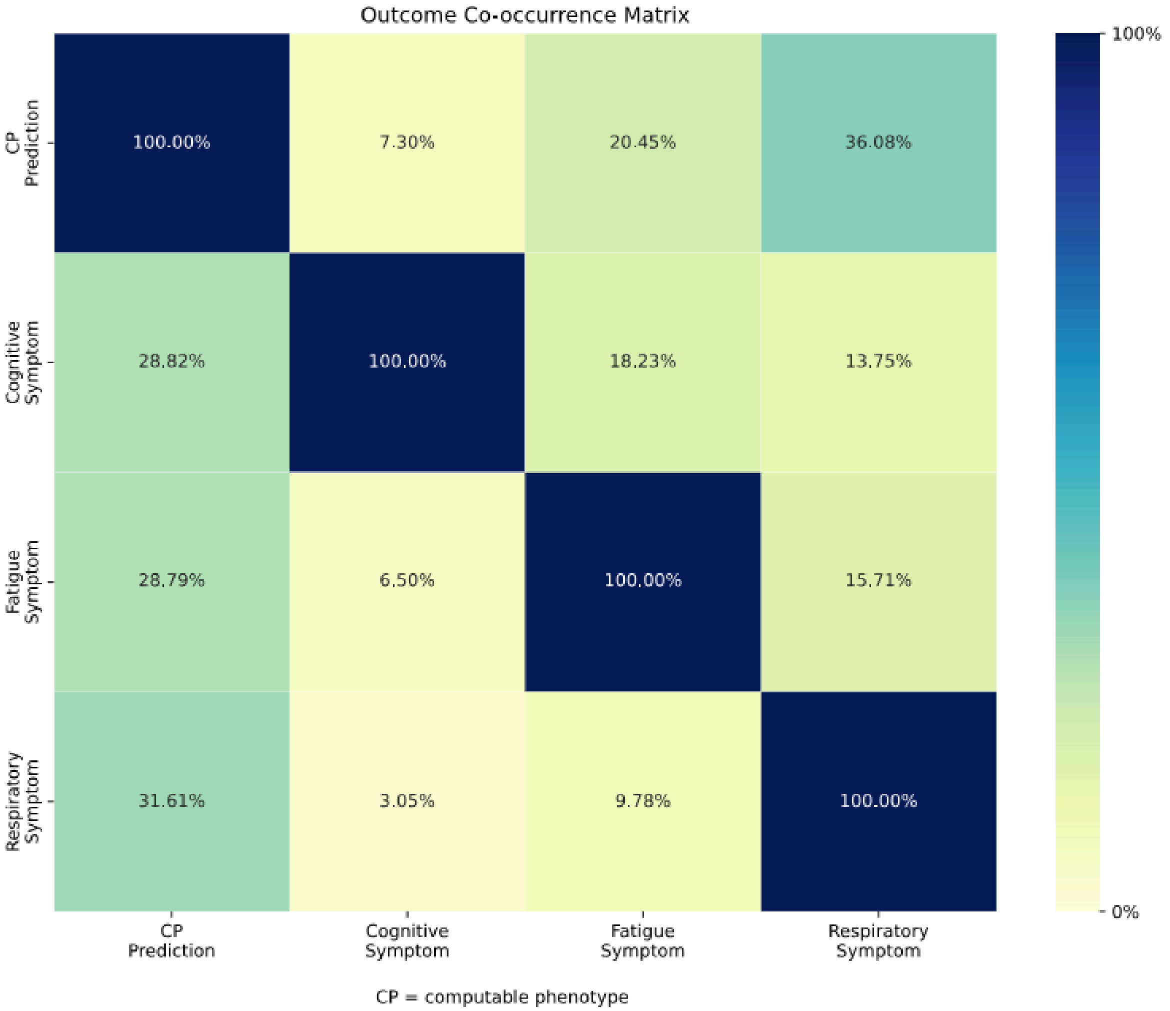

**S1 Figure B: Cumulative incidence of PASC in Paxlovid treated vs. Non-Paxlovid-Treated patients by outcome measure; between 29-180 days; VA-like subanalysis**

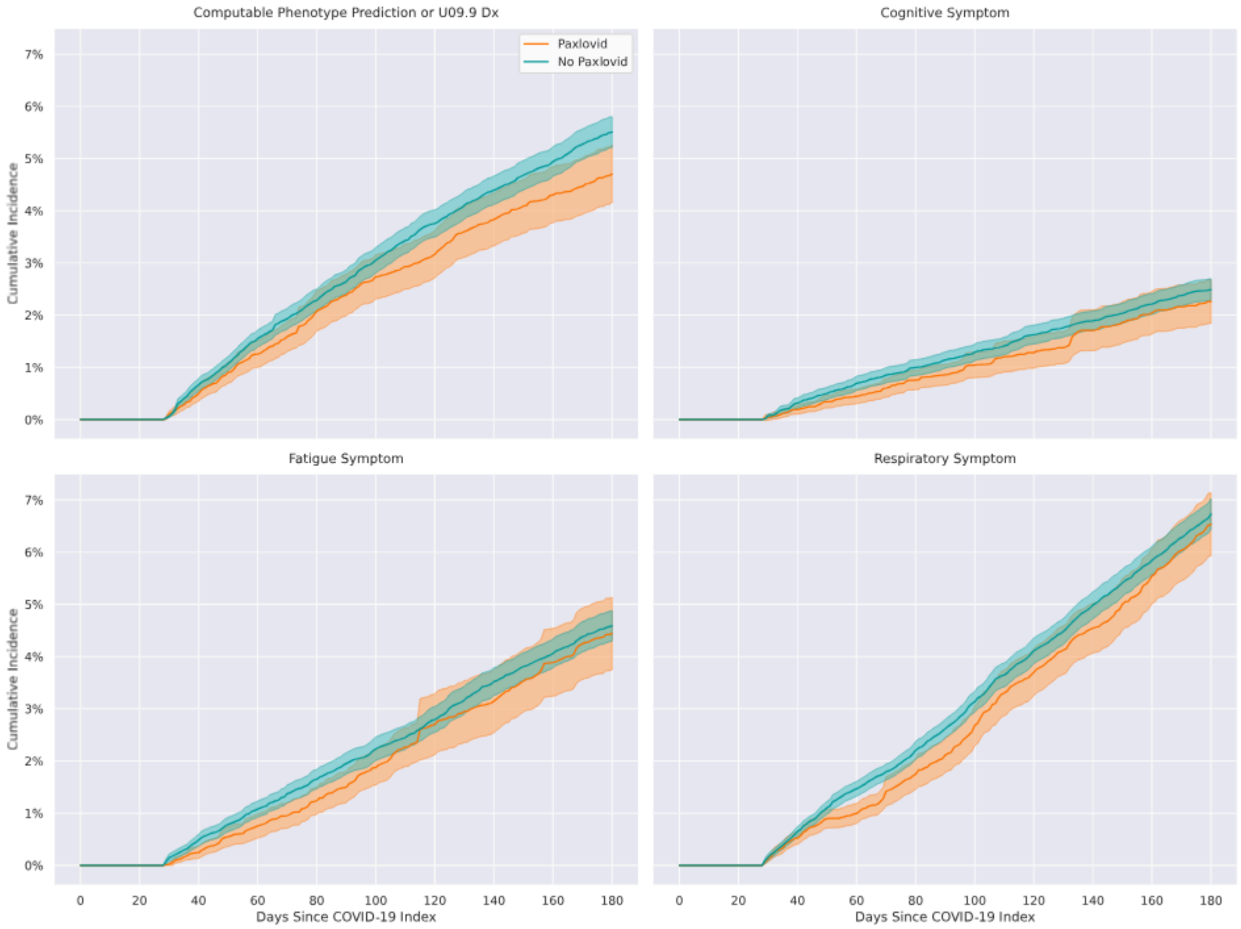

**S1 Figure C:** Cumulative incidence of PASC in Paxlovid treated vs. Non-Paxlovid-Treated patients by predicted outcome from CP model with threshold of 0.9 or U09.9, additionally adjusted for vaccination status and among data partners meeting vaccination data quality criteria

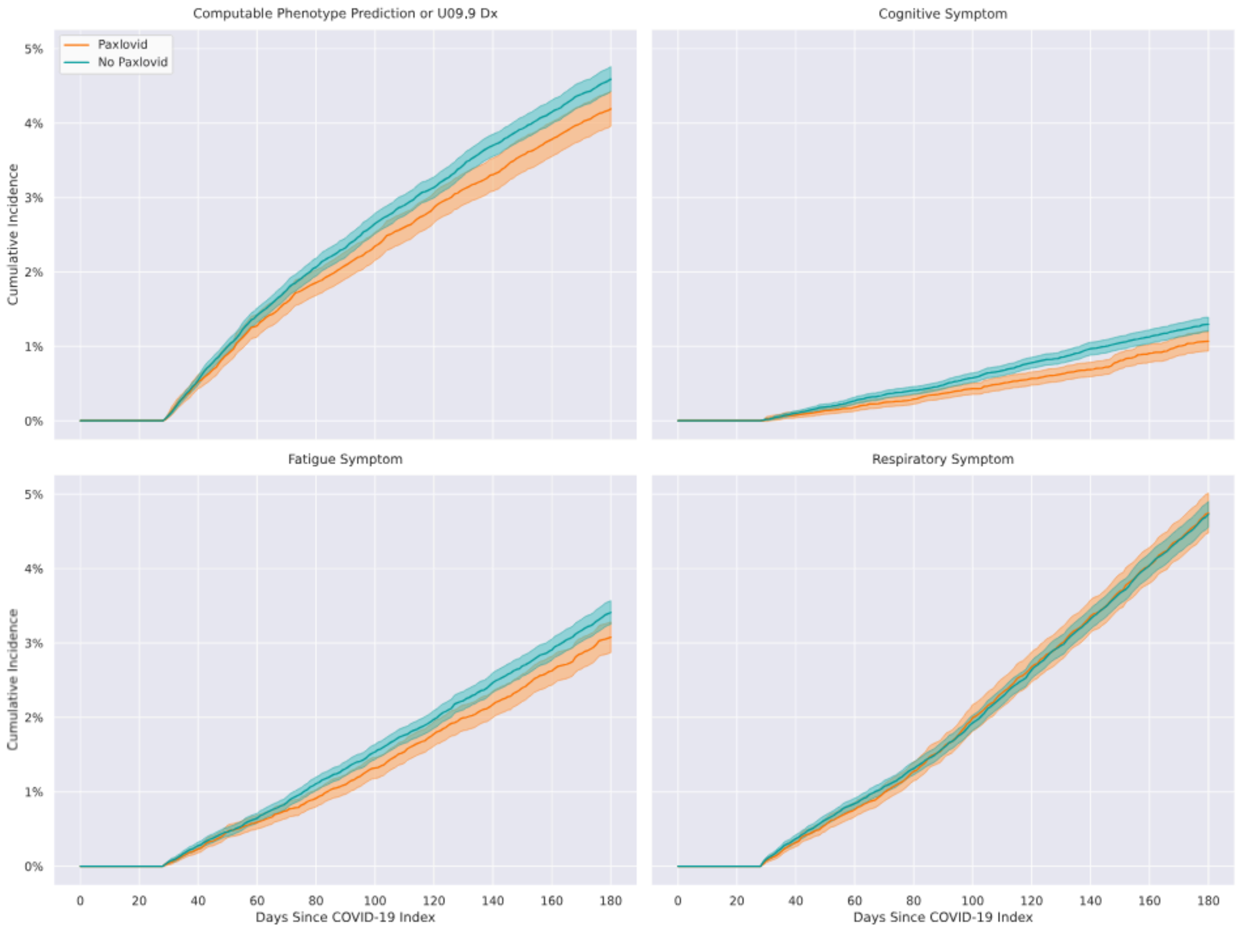

**S1 Figure D: Causal diagram used to inform covariate selection. Treatment is shown in green; outcome is shown in orange; observed covariates are shown in gray; unobserved covariates are shown in pink. Note that this diagram only shows the relationships relevant to this study. Covariates may have other causes that are omitted for clarity.**

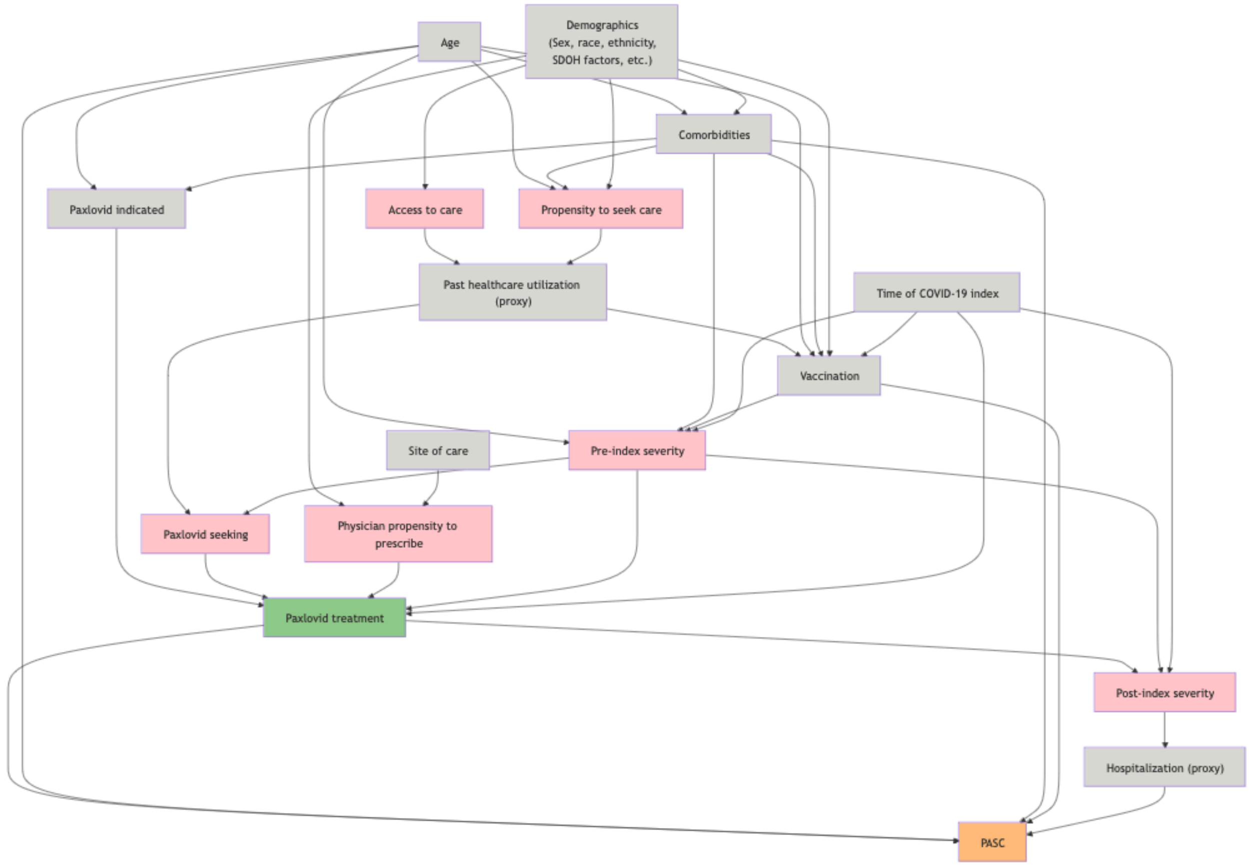

**S1 Table A:** ICD-10 codes used to define Global Burden of Disease symptom clusters[5]

| **ICD-10 Code** | **ICD-10 Code Description** | **Symptom Cluster** |
|---|---|---|
| R404 | Transient alteration of awareness | Cognitive |
| R410 | Disorientation unspecified | Cognitive |
| R411 | Anterograde amnesia | Cognitive |
| R412 | Retrograde amnesia | Cognitive |
| R413 | Other amnesia | Cognitive |
| R4182 | Altered mental status unspecified | Cognitive |
| R41840 | Attention and concentration deficit | Cognitive |
| R41841 | Cognitive communication deficit | Cognitive |
| R4189 | Other symptoms and signs involving cognitive functions and awareness | Cognitive |
| R419 | Unspecified symptoms and signs involving cognitive functions and awareness | Cognitive |
| R531 | Weakness | Fatigue |
| R5381 | Other malaise | Fatigue |
| R5382 | Chronic fatigue unspecified | Fatigue |
| R5383 | Other fatigue | Fatigue |
| J9610 | Chronic respiratory failure unspecified whether with hypoxia or hypercapnia | Respiratory |
| J9611 | Chronic respiratory failure with hypoxia | Respiratory |
| J9612 | Chronic respiratory failure with hypercapnia | Respiratory |
| J9620 | Acute and chronic respiratory failure unspecified whether with hypoxia or hypercapnia | Respiratory |
| J9621 | Acute and chronic respiratory failure with hypoxia | Respiratory |
| J9622 | Acute and chronic respiratory failure with hypercapnia | Respiratory |
| J9690 | Respiratory failure unspecified unspecified whether | Respiratory |

| | | |
|---|---|---|
| | with hypoxia or hypercapnia | |
| J9691 | Respiratory failure unspecified with hypoxia | Respiratory |
| J9692 | Respiratory failure unspecified with hypercapnia | Respiratory |
| J988 | Other specified respiratory disorders | Respiratory |
| J989 | Respiratory disorder unspecified | Respiratory |
| J99 | Respiratory disorders in diseases classified elsewhere | Respiratory |
| R05 | Cough | Respiratory |
| R0600 | Dyspnea unspecified | Respiratory |
| R0602 | Shortness of breath | Respiratory |
| R0603 | Acute respiratory distress | Respiratory |
| R0609 | Other forms of dyspnea | Respiratory |
| R071 | Chest pain on breathing | Respiratory |
